# Heterogeneous Associations of Latency Duration with Neonatal Outcomes After Preterm Prelabor Rupture of Membranes: A Nationwide Cohort Study

**DOI:** 10.64898/2026.09.07.26361886

**Authors:** Chaeyoon Shin, Jinyun Kim, Se In Sung, Yoonjung Yoonie Joo

## Abstract

**Importance:** Expectant management for preterm prelabor rupture of membranes (PPROM) is guided primarily by gestational age (GA), yet whether the association between latency duration and neonatal outcomes varies among pregnancies remains uncertain.

**Objective:** To characterize heterogeneous associations between latency duration and neonatal outcomes after PPROM and identify maternal and fetal characteristics associated with the largest estimated benefit.

**Design:** Nationwide registry-based retrospective cohort study using restricted cubic spline regression and causal forest analysis. Data were analyzed from July 2025 to July 2026.

**Setting:** The Korean Neonatal Network (KNN), a nationwide registry of infants born before 32 weeks’ gestation or with a birth weight less than 1,500g, from 2013 to 2023.

**Participants:** A total of 7,057 neonates born after PPROM were included.

**Exposures:** Latency duration, defined as the number of days from PPROM diagnosis to delivery (0-118 days).

**Main Outcomes and Measures:** Neonatal Health Index (NHI), an ordinal composite from 0 (death) to 8 (survival free of 7 major neonatal morbidities: intraventricular hemorrhage grades 3- 4, periventricular leukomalacia, retinopathy of prematurity stage ≥3, bronchopulmonary dysplasia, pulmonary hypertension, necrotizing enterocolitis stage ≥2, and sepsis).

**Results:** Among 7,057 neonates (median [IQR] maternal age, 34.0 [31.0-36.0] years; GA at PPROM, 27.0 [24.0-29.0] weeks; latency duration, 2.0 [0.0-8.0] days), longer latency was associated with better neonatal health, reflected by higher NHI (n = 6,872), in a non-linear, GA-dependent pattern (overall *P* < .001; nonlinearity *P* < .001). Causal forest analysis estimated an average partial effect of a 0.078-point increase in NHI per additional day of latency (*P* < .001), with significant heterogeneity across individuals (*P* < .001). GA at PPROM, oligohydramnios, and maternal age were the strongest modifiers; neonates with GA at PPROM of 26 weeks or less, no oligohydramnios, and maternal age younger than 30 years had an estimated treatment effect 28.0% greater than the rest of the cohort (0.121 vs 0.094 NHI points per latency-day).

**Conclusions and Relevance:** Associations between neonatal outcomes and prolonged latency after PPROM were positive overall but heterogeneous in magnitude, varying by GA at PPROM, oligohydramnios status, and maternal age. These findings support tailoring expectant management after PPROM according to individual patient characteristics rather than a uniform, GA-based strategy.

**Key points:** *Question:* Does the association between latency duration and neonatal outcomes after preterm prelabor rupture of membranes (PPROM) differ across pregnancies, and which characteristics identify the largest estimated benefit?

*Findings:* Among 7,057 neonates born after PPROM, longer latency was not uniformly associated with neonatal outcomes across pregnancies. The most favorable associations were observed in patients with earlier gestational age at PPROM (≤26 weeks), no oligohydramnios, and younger maternal age (<30 years).

*Meaning:* The neonatal benefit associated with longer latency after PPROM is concentrated in identifiable patient subgroups, which may refine and complement current gestational age-based expectant management.

## Introduction

Preterm birth affects approximately 13.4 million neonates annually and remains the leading cause of death in children younger than 5 years worldwide.^1^ Preterm prelabor rupture of membranes (PPROM) accounts for approximately 25% of preterm births,^2,3^ and neonates born following PPROM face elevated risks of periventricular leukomalacia (PVL), retinopathy of prematurity (ROP), bronchopulmonary dysplasia (BPD), necrotizing enterocolitis (NEC), and other major morbidities.^3^ Current guidelines recommend expectant management for PPROM before 34 weeks of gestation in the absence of maternal or fetal contraindications, balancing the maturational benefit of longer latency against the competing risks of intra-amniotic infection, placental abruption, and fetal compromise.^4^

However, these guidelines rely primarily on gestational age (GA) and indications for delivery, while providing limited evidence on whether the association between latency duration and neonatal outcomes varies according to baseline maternal and pregnancy characteristics.^4^ Furthermore, prior studies have typically categorized latency duration using fixed thresholds (e.g., <7 vs ≥7 days),^5–7^ estimating population-average associations with limited insight into which patients are most likely to benefit from longer latency. Identifying such patients requires moving beyond averaged, threshold-based summaries toward approaches that model nonlinear associations between latency duration and neonatal outcomes and estimate heterogeneity across clinically relevant baseline characteristics of each individual.

Causal forest is a machine learning-based causal inference approach that estimates individual-level treatment effects through data-adaptive recursive splitting and identifies factors driving heterogeneity without pre-specified interaction terms or functional-form assumptions.^8^ Unlike conventional subgroup analyses, which require potential effect modifiers a priori and may overlook complex interactions among correlated clinical characteristics, causal forest identifies patterns of treatment-effect heterogeneity directly from the data.^9^ By flexibly modeling nonlinearities and high-order interactions, this approach extends conventional regression beyond population-average estimates by generating patient-specific conditional estimates of how the association varies across baseline clinical profiles.^8,10^ The approach has been applied across several clinical domains, including obstetrics,^11^ critical care,^12^ and psychiatric research,^13–15^ yet its application to perinatal outcomes, and to PPROM in particular, remains unexplored.

Using data from the Korean Neonatal Network (KNN),^16^ a nationwide multicenter registry of very low birth weight infants or infants born before 32 weeks’ gestation, we investigated whether the association between latency duration after PPROM and neonatal outcomes was uniform across pregnancies or varied according to maternal and pregnancy characteristics at PPROM. Because PPROM before 34 weeks carries a high risk of very preterm birth, the KNN provides a clinically relevant population for evaluating GA-dependent neonatal morbidity, with outcomes prospectively collected using standardized definitions across centers. We combined restricted cubic spline (RCS) regression, which characterized the overall nonlinear latency– outcome relationship, with causal forest analysis, which estimated how that relationship differed across pregnancies and identified the characteristics driving those differences. Together, we aim to delineate clinical profiles for which longer latency showed the most favorable estimated associations with neonatal outcomes.

## Methods

### Study Design and Population

This comparative effectiveness, retrospective cohort study used data from the KNN, a nationwide registry of neonates born before 32 weeks of gestation or with a birth weight less than 1,500g across 77 institutions in South Korea.^16^ We included neonates registered from 2013 to 2023 who were born following PPROM, defined as spontaneous rupture of membranes before the onset of labor at less than 37 weeks’ gestation, and for whom latency duration could be calculated. Neonates were excluded if PPROM status was uncertain, if latency records were missing such that GA at PPROM could not be derived, or if latency exceeded 120 days, which was considered a data entry error, resulting in a final analytic sample of 7,057 neonates (**Figure S1**). The institutional review board at each participating NICU and the KNN data management committee approved the study, with a waiver of informed consent given the registry-based design. The study followed the International Society for Pharmacoeconomics and Outcome Research (ISPOR) Good Research Practices for nonrandomized comparative effectiveness research using secondary data sources^17^ and the Strengthening the Reporting of Observational Studies in Epidemiology (STROBE) guideline for cohort studies.^18^

### Exposures and Outcomes

Latency duration was defined as the number of days from PPROM diagnosis to delivery (0-118 days), calculated from PPROM diagnosis and delivery dates when both dates were available and otherwise from the directly recorded latency duration. A latency of 0 indicated delivery within 24 hours after diagnosis. GA at PPROM was derived by subtracting the latency duration from GA at delivery.

The primary outcome was the Neonatal Health Index (NHI), an ordinal composite measure ranging from 0 (death) to 8 (no major morbidity), with higher scores indicating better neonatal health. Among survivors, the score was calculated as 8 minus the number of major neonatal morbidities present, including grade 3 or 4 intraventricular hemorrhage (IVH), periventricular leukomalacia (PVL), retinopathy of prematurity stage ≥3 (ROP), bronchopulmonary dysplasia (BPD), pulmonary hypertension (PH), necrotizing enterocolitis stage ≥2 (NEC), and neonatal sepsis, as recorded in the KNN before discharge. Neonatal death and each morbidity were analyzed individually as secondary outcomes, modeled as binary variables and reverse-coded in causal forest analyses (case=0, control=1) so that higher values represented better neonatal health.

The NHI was specified a priori to preserve neonatal death as the most severe outcome while assigning equal weight to each major morbidity among survivors. The index was informed by prior PPROM outcome studies and earlier composite outcome measures that jointly captured neonatal death and severe morbidity.^19,20^ Given the lack of a universally adopted composite outcome for PPROM-related prognosis,^21^ the index was restricted to morbidities with established severity thresholds.^22^

NHI analyses included 6,872 neonates with complete data on death and all component morbidities; analyses of secondary outcomes used all available nonmissing outcome data.

### Covariates and Effect Modifiers

Pre-exposure maternal and perinatal characteristics were selected a priori based on clinical relevance and their potential association with both latency duration and neonatal outcomes. The covariate set was restricted to variables measured before or at the time of PPROM diagnosis to avoid adjustment for mediators or post-exposure factors, including GA at delivery, antenatal corticosteroid use, and histologic chorioamnionitis. Maternal covariates included age at delivery, parity (primiparity or multiparity), mode of conception (spontaneous or in vitro fertilization), diabetes (gestational or overt), hypertension (pregnancy-induced or chronic), and oligohydramnios. Oligohydramnios was coded as a binary variable (present or absent), with polyhydramnios and normal amniotic fluid volume as the reference category, given its limited prognostic relevance in PPROM compared with oligohydramnios.^23^ Perinatal covariates included fetal sex, plurality (singleton or multiple), GA at PPROM, and size for GA (small [SGA], appropriate [AGA], or large [LGA]) defined by the Korean birth weight percentile reference.^24,25^ Size for GA was derived from birth weight, served as a proxy for fetal growth in the absence of antenatal estimated fetal weight. Oligohydramnios was the only covariate with missing data, which was imputed using MICE algorithm^26^ (miceforest package v6.0.3, Python v3.11.0).

### Statistical Analysis

RCS regression was used to characterize the nonlinear association between latency duration and neonatal outcomes. Candidate knots were set at 3, 7, 14, 21, and 28 days based on prior studies^6,7,27^ and the observed data distribution, omitting knots outside each subgroup’s observed latency range. RCS terms were incorporated into a proportional odds regression model with robust variance estimation for NHI (treated as an ordinal count of health, 0–8) and logistic regression models for the binary secondary outcomes.^28^ Models were fitted in the full cohort with GA at PPROM as a covariate and separately within four GA at PPROM subgroups (<24, 24- 25+6/7, 26-27+6/7, and ≥28 weeks), each adjusted for the covariates described above, allowing the shape of the association to vary across GA at PPROM strata. Statistical significance of the overall association and nonlinearity were assessed using Wald tests. Average marginal predictions were computed across the observed covariate distribution with 95% CIs from 500 bootstrap resamples and truncated at the 99th percentile of latency duration distribution within each GA at PPROM subgroup to avoid unstable estimates. All analyses were performed using rms package (v8.0.0)^29^ in R (v4.4.3).

Continuous variables are reported as median [IQR] and compared across GA at PPROM subgroups with Kruskal-Wallis test; categorical variables are reported as frequency (%) and compared with chi-square tests.

### Causal Forest Analysis

Treatment effects of latency duration, modeled as a continuous exposure, were estimated using causal forests within the generalized random forest (GRF) framework.^8^ The average partial effect (APE) represents the average marginal change in the outcome per additional day of latency, and the conditional average treatment effect (CATE) is the expected change conditional on baseline characteristics. The APE is an average slope across the observed latency range and does not imply a constant per-day effect. Individual-level CATE estimates are hereafter referred to as individual treatment effects (ITEs). Primary analyses were conducted using *grf* package (v2.5.0) in R, with *EconML* package (v0.16.0) in Python used to corroborate APE estimates and conduct supplementary analyses. Model fit was assessed with the *grf* calibration test, which evaluates how well the predicted APE and ITEs reflect the observed latency–outcome associations. Sensitivity to unmeasured confounding was evaluated using the omitted variable bias (OVB) framework implemented in *EconML*. Detailed descriptions are provided in **Supplementary Methods**.

To assess treatment effect heterogeneity, patients were stratified into ITE-based quintiles. Group average treatment effects (GATEs) were estimated within each quintile.^13,15,30–32^ Quintile- to-quintile differences (Q2–Q5 vs Q1) were tested using two-sided Wald z-tests. Quartile- and decile-based groupings were additionally examined. Baseline covariates of the lowest- and highest-benefit pregnancies (Q1 vs Q5) were compared using Student’s t-tests for continuous variables, and Fisher’s exact or chi-squared tests for categorical variables with standardized mean differences (SMDs) calculated.

Variable importance identified covariates contributing most to treatment-effect heterogeneity learned by the causal forest. Best linear projection (BLP), regressing doubly robust scores on all covariates, provided a linear summary of each covariate’s association with ITE variation conditional on others.

### Interpreter Tree and Subgroup Analysis

To identify clinically interpretable subgroups, we fit a shallow regression tree to ITEs using the *SingleTreeCateInterpreter* function in *EconML*. The algorithm splits on covariates that maximize between-node differences in CATEs, each terminal node reporting the APE for its patients. Interpreter trees were derived under multiple hyperparameter settings (maximum depth = 2–4; minimum leaf size = 500, 1,000, 1,500) to verify structural stability. Larger minimum leaf sizes (≥ 2,000) were not considered as they produced trees dominated by a single variable (GA at PPROM), limiting exploration of heterogeneity. Variables recurring as splitting nodes across the majority of interpreter trees were considered candidate effect modifiers, and the tree with the most frequently recurring structure was selected as representative. We additionally constructed heatmaps of *grf*-derived ITEs across prespecified clinical strata of these variables.

### Sensitivity Analysis

To account for within-pregnancy correlation among neonates from multiple gestations,^33,34^ we repeated the RCS and causal forest analyses using a proxy pregnancy identifier, constructed from identical birth date, maternal age, parity, and gestational age at delivery. Pregnancy-level clustering was incorporated into both analyses.^35^ We also repeated the NHI causal forest analysis in pregnancies with latency of 30 days or less (n=6,575) to assess robustness after excluding observations with extreme latency values and limited covariate overlap (**Supplementary Methods**).

## Results

### Study Population

The study cohort comprised 7,057 neonates. Participants were stratified by GA at PPROM into four subgroups: <24 weeks (n = 1,293; 18.3%), 24-25+6 weeks (n = 1,272; 18.0%), 26-27+6 weeks (n = 1,445; 20.5%), and ≥28 weeks (n = 3,047; 43.2%). Median values were 34.0 years [IQR, 31.0-36.0] for maternal age, 27.0 weeks [IQR, 24.0-29.0] for GA at PPROM, and 2.0 days [IQR, 0.0-8.0] for latency. Earlier GA at PPROM was associated with longer and more broadly distributed latency (median 19.0 days in the <24-week group vs 0.0 days in the ≥28-week group; *P* < .001) (**Figure 1A**).

**Figure 1.**
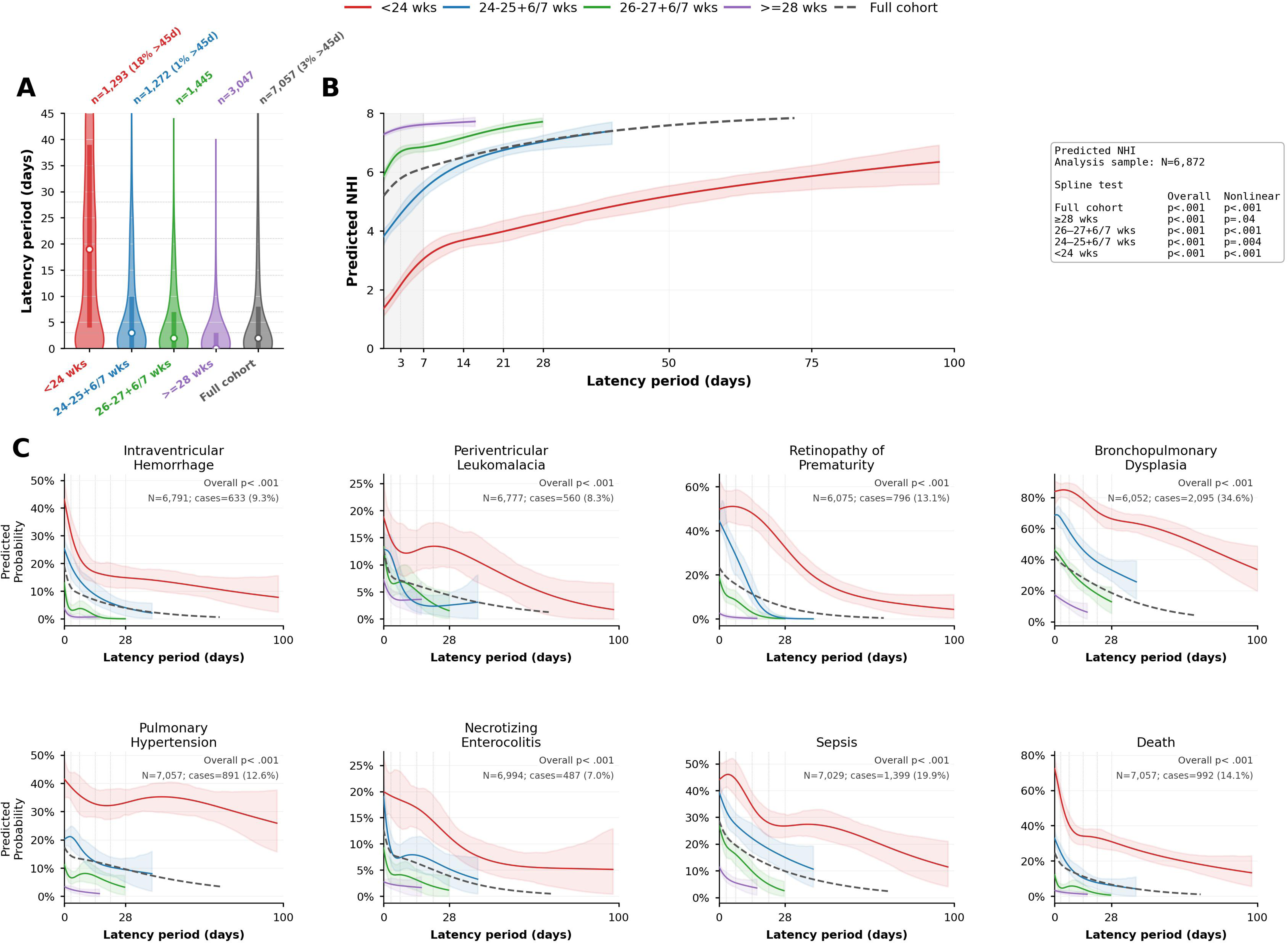
Latency distribution and dose-response relationship between latency period and neonatal outcomes according to gestational age (GA) at preterm prelabor rupture of membranes (PPROM). **A**, Distribution of latency periods by GA at PPROM stratum (<24, 24-25+6/7, 26-27+6/7, and ≥28 weeks), shown as vertical violin plots (white dot, median; thick bar, interquartile range). The y- axis is truncated at 45 days for display; dotted horizontal lines mark the fixed knot locations (3, 7, 14, 21, and 28 days). For strata with observations beyond 45 days, the percentage of values exceeding the cutoff is noted above the corresponding violin. The analysis included the full cohort (N=7,057). **B,** Covariated-adjusted dose-response association between latency period and the Neonatal Health Index (NHI). The analysis included infants with complete information required to calculate the NHI (n = 6,872). Fixed knots were placed at 3, 7, 14, 21, and 28 days and are indicated by vertical gray dashed lines. The gray dashed curve represents the full-cohort model and the colored solid curves represent the subgroup-specific models. Shaded areas indicate 95% CIs. Curves are displayed up to the 99th percentile of the observed latency distribution within each subgroup. Overall and nonlinearity P values are summarized in the table. **C**, Covariate-adjusted dose-response associations between latency period and secondary outcomes (neonatal death and morbidities). Outcome-specific analytic sample sizes and case counts, reflecting differences in outcome availability, are shown above each panel.

Death before hospital discharge occurred in 14.1% of neonates overall, ranging from 2.9% in the ≥28-week group to 39.1% in the <24-week group (Table 1). The most prevalent morbidities were BPD (29.7%), sepsis (19.8%), and PH (12.6%); all rates differed significantly across GA at PPROM subgroups (*P* < .001). Among the 6,872 of 7,057 neonates (97.4%) with complete outcome data, the median NHI was 7.0 [IQR, 6.0–8.0] overall and 5.0 [IQR, 0.0-6.0] in the <24- week group.

**Table 1.** Demographics of the study cohort.

|  |  |  | GA at PPRM |  |  |  |  |
| --- | --- | --- | --- | --- | --- | --- | --- |
|  |  | Overall | <24 weeks | 24-25+6/7 weeks | 26-27+6/7 weeks | >=28 weeks | *P*-value |
| n |  | 7057 | 1293 | 1272 | 1445 | 3047 |  |
| Maternal age (years), median [Q1,Q3] |  | 34.0<br>[31.0,36.0] | 34.0<br>[31.0,36.0] | 34.0<br>[31.0,36.0] | 34.0<br>[31.0,37.0] | 33.0<br>[31.0,36.0] | .005 |
| GA at PPRM (weeks), median [Q1,Q3] |  | 27.0<br>[24.0,29.0] | 22.0<br>[20.0,23.0] | 25.0<br>[24.0,25.0] | 27.0<br>[26.0,27.0] | 29.0<br>[28.0,30.0] | <.001 |
| Latency duration (days), median [Q1,Q3] |  | 2.0<br>[0.0,8.0] | 19.0<br>[4.0,39.0] | 3.0<br>[0.0,10.0] | 2.0<br>[0.0,7.0] | 0.0<br>[0.0,3.0] | <.001 |
| Fetal sex (Male), n (%) |  | 3639<br>(51.6) | 682<br>(52.7) | 678<br>(53.3) | 792<br>(54.8) | 1487<br>(48.8) | <.001 |
| Multiple gestation, n (%) |  | 2646<br>(37.5) | 485<br>(37.5) | 356<br>(28.0) | 387<br>(26.8) | 1418<br>(46.5) | <.001 |
| IVF conception, n (%) |  | 2135<br>(30.3) | 462<br>(35.7) | 357<br>(28.1) | 357<br>(24.7) | 959<br>(31.5) | <.001 |
| GDM or overt DM, n (%) |  | 857<br>(12.1) | 94 (7.3) | 120 (9.4) | 179<br>(12.4) | 464<br>(15.2) | <.001 |
| Chronic HTN or PIH, n (%) |  | 366 (5.2) | 50 (3.9) | 40 (3.1) | 60 (4.2) | 216 (7.1) | <.001 |
| Oligohydramnios, n (%) |  | 1499<br>(21.2) | 487<br>(37.7) | 295<br>(23.2) | 287<br>(19.9) | 430<br>(14.1) | <.001 |
| Primiparity, n (%) |  | 4516<br>(64.0) | 836<br>(64.7) | 758<br>(59.6) | 843<br>(58.3) | 2079<br>(68.2) | <.001 |
| Size for GA, n (%) | **SGA** | 461 (6.5) | 49 (3.8) | 30 (2.4) | 31 (2.1) | 351 (11.5) | <.001 |
|  | **AGA** | 6495 (92.0) | 1212 (93.7) | 1209 (95.0) | 1385 (95.8) | 2689 (88.3) |  |
|  | **LGA** | 101 (1.4) | 32 (2.5) | 33 (2.6) | 29 (2.0) | 7 (0.2) |  |
| IVH, n (%) |  | 633 (9.0) | 240 (18.6) | 217 (17.1) | 99 (6.9) | 77 (2.5) | <.001 |
| PVL, n (%) |  | 560 (7.9) | 141 (10.9) | 112 (8.8) | 127 (8.8) | 180 (5.9) | <.001 |
| ROP, n (%) |  | 796 (11.3) | 272 (21.0) | 304 (23.9) | 163 (11.3) | 57 (1.9) | <.001 |
| BPD, n (%) |  | 2095 (29.7) | 553 (42.8) | 568 (44.7) | 513 (35.5) | 461 (15.1) | <.001 |
| PH, n (%) |  | 891 (12.6) | 455 (35.2) | 225 (17.7) | 122 (8.4) | 89 (2.9) | <.001 |
| NEC, n (%) |  | 487 (6.9) | 172 (13.3) | 149 (11.7) | 89 (6.2) | 77 (2.5) | <.001 |
| Sepsis, n (%) |  | 1399 (19.8) | 427 (33.0) | 392 (30.8) | 291 (20.1) | 289 (9.5) | <.001 |
| Death, n (%) |  | 992 (14.1) | 506 (39.1) | 288 (22.6) | 111 (7.7) | 87 (2.9) | <.001 |
| NHI (0-8), median [Q1,Q3] |  | 7.0 [6.0,8.0] | 5.0 [0.0,6.0] | 6.0 [3.0,7.0] | 7.0 [6.0,8.0] | 8.0 [7.0,8.0] | <.001 |

GA, Gestational Age; PPRM, Preterm Prelabor Rupture of Membranes; IVF, In-Vitro Fertilization; GDM, Gestational Diabetes Mellitus; DM, Diabetes Mellitus; HTN, Hypertension; PIH, Pregnancy-Induced Hypertension; IVH, Intraventricular hemorrhage; PVL, Periventricular leukomalacia; ROP, Retinopathy of prematurity; Bronchopulmonary dysplasia, BPD; PH, Pulmonary hypertension; NEC, Necrotizing enterocolitis; SGA, small for GA; AGA, appropriate for GA; LGA, large for GA; NHI, Neonatal Health Index

### Restricted Cubic Spline Analysis

In the covariate-adjusted full-cohort model, both latency duration and GA at PPROM were associated with NHI (*P* < .001). Associations were nonlinear for NHI, IVH, PVL, NEC, sepsis, and death (nonlinearity *P* ≤ .007), but not for ROP, BPD, or PH (**Table S1**). Predicted NHI increased most steeply during the first week of latency, from 5.19 (95% CI, 5.09-5.28) at day 0 to 6.12 (95% CI, 6.04-6.19) at day 7, and more gradually thereafter, reaching 7.06 (95% CI, 7.00- 7.13) by day 28 (**Figure 1B, Table S3)**.

In GA at PPROM subgroup analyses, latency remained associated with NHI across all GA strata (*P* < .001), with evidence of nonlinearity in each stratum (*P* = .04 to < .001) (**Table S2**). The magnitude of the association was greatest at earlier GA at PPROM and diminished with advancing GA (**Figure 1B, Table S3**). The <24-week group showed the largest increase in predicted NHI with longer latency, from 1.36 (95% CI, 1.10-1.66) at day 0 to 6.33 (95% CI, 5.59-6.90) at day 97, with the steepest rise occurring during the first week. In the 24–25+6-week and 26–27+6-week groups, predicted NHI also increased with longer latency, but the magnitude of increase was progressively smaller. The ≥28-week group showed the highest predicted NHI at day 0, 7.28 (95% CI, 7.21-7.36), and the smallest increase with longer latency.

Latency was associated with all secondary outcomes in the full-cohort model (*P* < .001) **(Figure 1C, Table S1)**. In subgroup analyses, associations were not statistically significant for PH in the <24-week (*P* = .15) or ≥28-week (*P* = .1) groups, or NEC (*P* = .6) or death (*P* = .17) in the ≥28-week group (**Table S2**).

### Causal Forest Analysis

The causal forest estimated that each additional day of latency was associated with an average increase of 0.078 NHI points (*P* < .001), with all ITEs positive and spanning an approximately 2-fold range, from 0.065 to 0.144. Calibration diagnostics indicated good model fit (βATE = 0.908, *P* < .001). The heterogeneity calibration coefficient was βITE = 1.709 (*P* < .001), indicating that the model detected treatment effect heterogeneity but likely underestimated its magnitude. An independent implementation in EconML showed a similar point estimate (APE = 0.081; *P* = .02). In the sensitivity analysis using the OVB framework, the bias-adjusted APE (θ) was 0.116 (partially identified interval, 0.097-0.135; 95% CI, 0.088-0.144) (**Table S4**). The robustness value for θ was 0.269 and 0.249 for the 95% CI, indicating that an unobserved confounder would need to explain 26.9% of the residual variation in both latency and NHI to nullify the estimate to zero, and 24.9% to render it insignificant.

APEs and model fits of the secondary outcomes are presented in **Table S5**. All outcomes had significant and well-calibrated APEs (*P* < .001; NEC, *P* = .02). However, PH, PVL, and NEC showed unstable heterogeneity calibration coefficients, indicating that between-individual variation in estimated effects on these outcomes was not reliably captured by the model.

### Characterization of Treatment Effect Heterogeneity

Patients were stratified into quintiles of estimated ITEs, and group average treatment effects (GATEs) were estimated within each quintile. Quintiles were numbered from Q1, the fifth of pregnancies with the smallest predicted effect, through Q5, the largest. Quintile GATEs rose monotonically across quintiles, from 0.041 to 0.134 NHI points per additional latency day (both P < .001), consistent with the heterogeneity indicated by the calibration test (**Figure 2A**). The APEs of Q4 and Q5 differed significantly from Q1, indicating that detectable heterogeneity was possibly concentrated in the upper quintiles rather than spread evenly across the distribution (**Figure 2A, Table S6-S7**). Quartile- and decile-based analyses showed similar monotonic patterns (**Figure S2, Table S6).** Among secondary outcomes, quintile GATEs for PH, PVL, sepsis, and NEC (**Figure S3**) were not observed to increase across quintiles, with APEs of Q5 either non-significant or smaller than those of lower quintiles. This aligned with the calibration test results, in which PH, PVL, and NEC had βITE estimates that differed substantially from 1 (**Table S5**).

**Figure 2.**
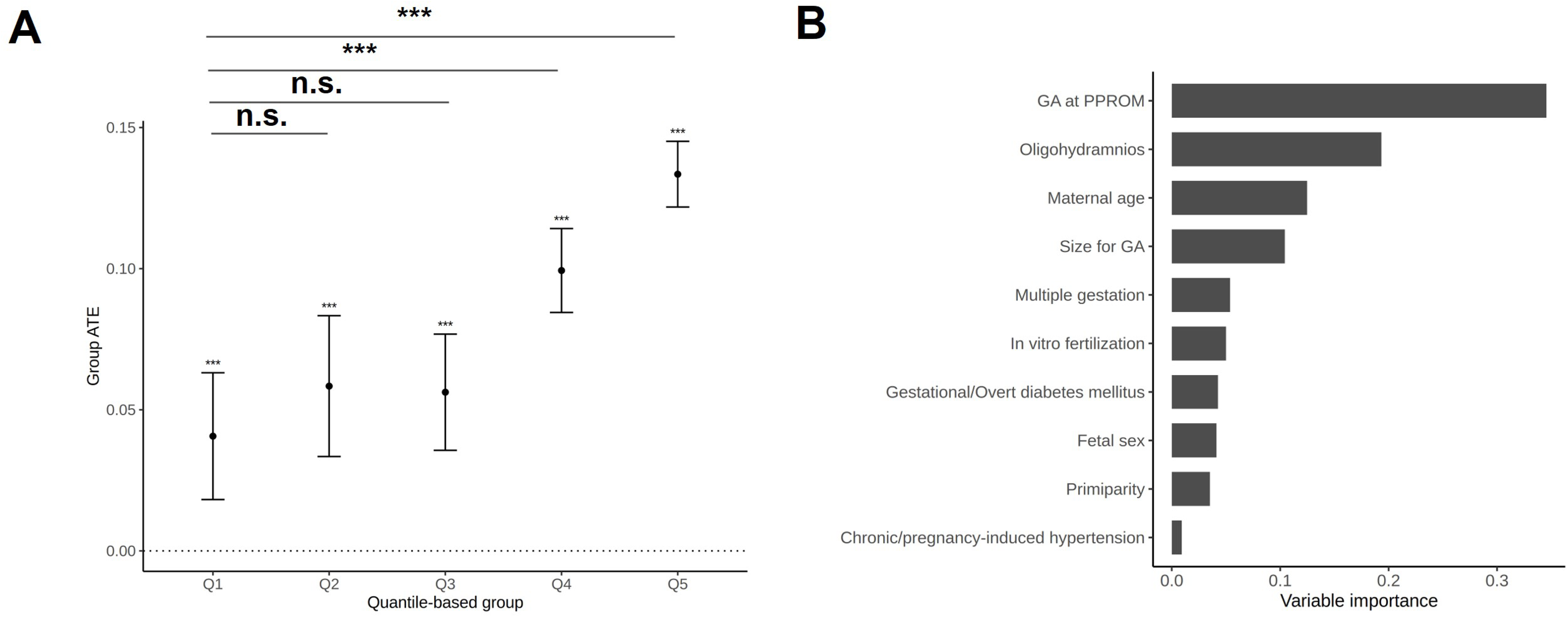
Identification of variables contributing to treatment effect heterogeneity. **A**, Quintile-based group average treatment effect (GATE) test. Patients were stratified into quintiles according to predicted individual treatment effects (ITEs), and the estimated GATE with 95% confidence intervals is shown for each quantile group. Asterisks indicate statistical significance of the GATE estimates, and horizontal bars denote pairwise comparisons between Q1 and the other quintiles. **B**, Variable importance from the causal forest fitted in *grf*. *** p < .001; ** p < .01; * p < .05

Baseline characteristics differed markedly between the lowest- and highest- benefit quintiles (Q1 vs Q5; **Table 2**). Relative to Q1, Q5 comprised pregnancies with younger maternal age, earlier GA at PPROM, lower prevalences of in vitro fertilization, gestational or overt diabetes mellitus, hypertension, oligohydramnios, and primiparity, and higher proportions of singleton pregnancies and male fetuses. Size for GA distribution also differed between groups. GA at PPROM showed the largest between-group separation (SMD, −3.29), followed by multiple gestation (SMD, −0.37) and oligohydramnios (SMD, −0.28), whereas hypertension (SMD, −0.02) and LGA (SMD, 0.02) showed minimal separation (**Table 2**).

**Table 2.** Covariate comparison between the low- (Q1) and high-benefit (Q5) group.

| Variable |  | Q1<br>(n=1375) | Q5<br>(n=1375) | p | Standardized<br>mean difference |
| --- | --- | --- | --- | --- | --- |
| Maternal age (mean (SD)) |  | 33.7 (2.8) | 33.0 (4.3) | <.001 | -0.13 |
| GA at PPRM (weeks)<br>(mean (SD)) |  | 29.1 (1.9) | 22.9 (1.9) | <.001 | -3.29 |
| Multiple gestation, n(%) |  | 970<br>(70.5%) | 481 (35.0%) | <.001 | -0.37 |
| IVF conception, n(%) |  | 800<br>(58.2%) | 409 (29.7%) | <.001 | -0.28 |
| GDM or overt DM, n(%) |  | 220<br>(16.0%) | 74 (5.4%) | <.001 | -0.11 |
| Chronic HTN or PIH, n(%) |  | 68 (4.9%) | 43 (3.1%) | <.001 | -0.02 |
| Oligohydramnios, n(%) |  | 459<br>(33.4%) | 4 (0.3%) | <.001 | -0.28 |
| Primiparity, n(%) |  | 1173<br>(85.3%) | 883 (64.2%) | <.001 | -0.24 |
| Fetal sex (Male), n(%) |  | 551<br>(40.1%) | 691(50.3%) | <.001 | -0.1 |
| Size for GA, n(%) | SGA | 199<br>(14.5%) | 22 (1.6%) | <.001 | -0.12 |
|  | AGA | 1167<br>(84.9%) | 1314<br>(95.6%) |  | 0.1 |
|  | LGA | 9 (0.7%) | 39 (2.9%) |  | 0.02 |

IVF, In-Vitro Fertilization; GDM, Gestational Diabetes Mellitus; DM, Diabetes Mellitus; HTN, Hypertension; PIH, Pregnancy-Induced Hypertension; SGA, small for GA; AGA, appropriate for GA; LGA, large for GA

Variable importance ranked GA at PPROM, oligohydramnios, and maternal age highest among the candidate modifiers (**Figure 2B, Table S8**). Results for secondary outcomes are shown in **Figure S4**. In the BLP, hypertension (β = 0.057), earlier GA at PPROM (β = −0.009 per week), and absence of oligohydramnios (β = −0.042) were associated with larger predicted effects (**Table S9**).

### Identification of High-Benefit Subgroup

With positive estimated effects in every pregnancy, interpreter trees were fit to characterize the covariate profile with the largest estimated effects. GA at PPROM (≤26.5 weeks) consistently emerged as the root node under every hyperparameter setting examined, identifying it as the covariate that best separated estimated effects at the first split (**Figure S5**). Subsequent splits most frequently involved oligohydramnios, maternal age, and multiple gestation. Splits on multiple gestation were not reproduced across hyperparameter settings and yielded imprecise CATE estimates, so interpretation focused on the three modifiers with stable estimates: GA at PPROM, oligohydramnios, and maternal age. In the representative tree (maximum depth = 3, minimum leaf node size = 1000), the largest CATE (0.13; 95% CI, 0.07-0.18) was observed in those with early GA at PPROM (≤26.5 weeks), no oligohydramnios, and younger maternal age (≤32.5 years) (**Figure 3A**). Heatmaps based on clinically defined strata and *grf*-derived ITEs showed patterns consistent with the interpreter tree (**Figure 3B**). The subgroup with GA at PPROM ≤26 weeks, no oligohydramnios, and maternal age <30 years had a higher CATE than the rest of the cohort (0.121 [n = 331] vs 0.094 [n = 6541]; difference, 0.027 NHI points per latency-day (95% CI, 0.0249- 0.0278; t = 35.57; P < .001), corresponding to a relative 28.0% higher CATE.

**Figure 3.**
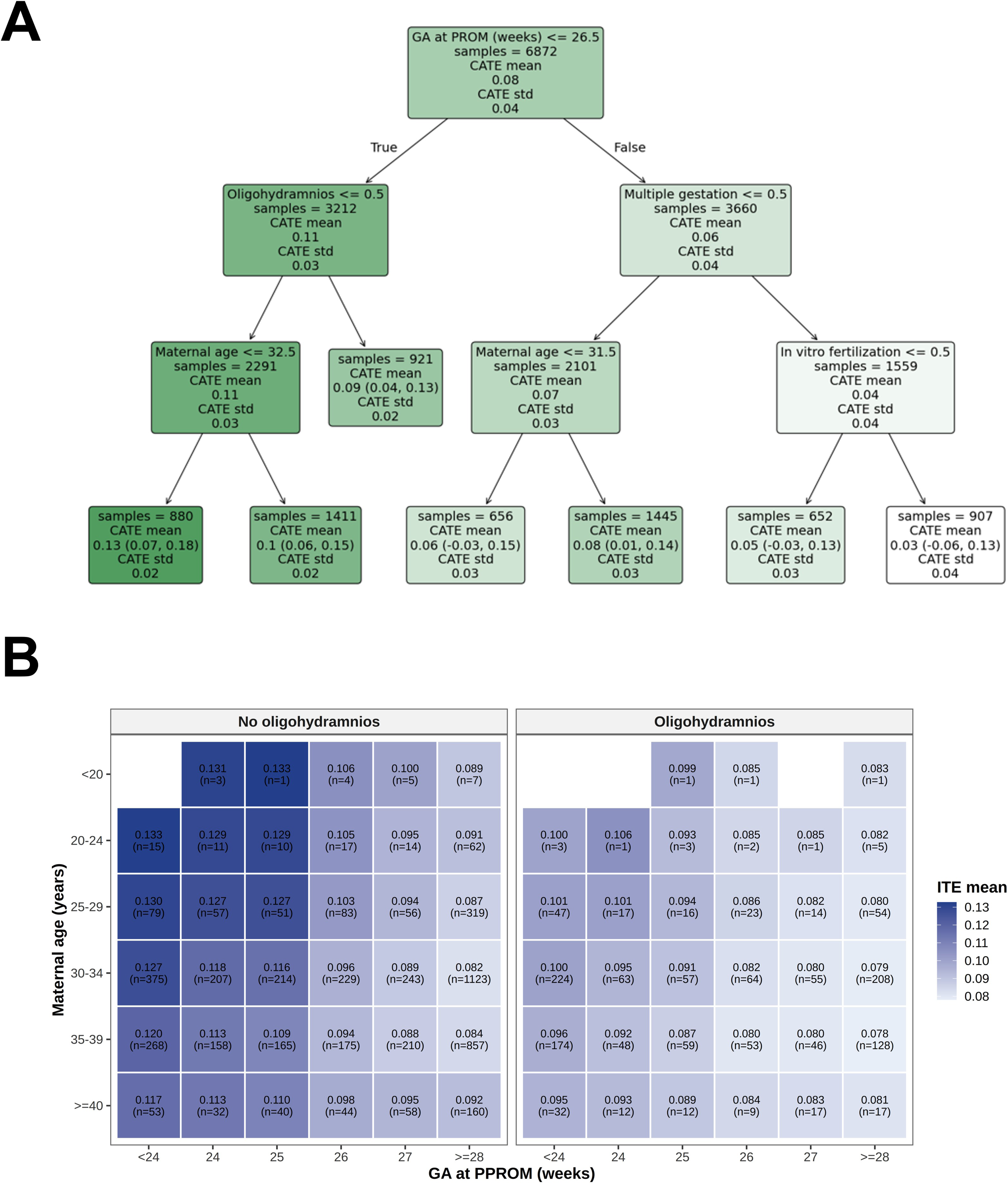
Identification of subgroups with greater benefit from prolonged latency. **A**, CATE (conditional average treatment effect) interpreter tree. Each node represents a split based on covariates, and terminal nodes display the estimated CATE, number of patients, and associated statistics (mean, standard deviation) for the corresponding subgroup. **B**, Heatmap of CATEs of latency period within subgroups divided by GA at PPROM, maternal age, and oligohydramnios status. Numbers inside each cell indicate the estimated CATE, which is the mean of the individual treatment effects (ITEs) within the subgroup, with the sample size in parentheses. Darker shading indicates larger estimated treatment effects.

### Sensitivity Analysis

Of the 7,057 neonates, 2,646 (37.5%) were from multiple gestations, yielding 780 clusters (742 twin pairs, 38 triplet sets; 1,598 neonates); the remaining multiples did not have identifiable co-twin within the registry. In RCS, predicted NHI point estimates were unchanged to two decimal places across all timepoints and GA subgroups, and confidence intervals differed only marginally in width (**Table S10**), indicating that within-cluster correlation did not materially influence the estimated associations between latency duration and neonatal outcomes. Similarly, the estimated APE remained unchanged (0.078, *P* < .001), with calibration metrics similar to those of the primary analysis (βATE = 0.892 and βITE = 1.903; both *P* < .001) in the causal forest.

Restricting the sample to pregnancies with latency ≤ 30 days (n = 6,575), the APE remained significant in both *grf* (0.087, *P* < .001) and *EconML* (0.093, *P* < .001) (**Table S11**) with good calibration (βATE = 0.903, *P* < .001; βITE = 1.278, *P* < .001). The quintile-based GATE test showed significant, monotonically increasing effects across quintiles (**Figure S6A, Table S12- S13**). Comparison of covariate distribution between the lowest and highest quintiles (Q1 vs. Q5) indicated significant differences for all covariates except maternal age (*P* = .09), and the prevalence patterns were consistent with the main analysis (**Table S14**). GA at PPROM (SMD = 5.17), followed by multiple gestation (SMD = 0.51), and primiparity (SMD = 0.30) were the most differential between the two groups. GA at PPROM, maternal age, and oligohydramnios showed the highest variable importance, coherent with the main analysis (**Figure S6B, Table S15**). BLP identified that having hypertension (β = 0.053) oligohydramnios (β = −0.042), and earlier GA at PPROM (β = −0.017) was significantly associated with ITE variation, reproducing the directions observed in the main analysis (**Table S16**).

## Discussion

In this nationwide cohort of 7,057 neonates born after PPROM, we conducted, to our knowledge, the first study to characterize how the association between latency duration and neonatal outcomes differs across pregnancies. The estimated neonatal benefit of longer latency varied approximately 2-fold across individuals, indicating substantial heterogeneity beyond the population-average association. GA at PPROM was the dominant modifier of the estimated benefit, extending prior evidence identifying it as a key determinant of both latency and perinatal outcomes,^3,27,36^ while oligohydramnios status and maternal age further distinguished pregnancies with larger estimated effects. The subgroup with the highest-benefit profile showed an estimated effect 28.0% greater than the remaining cohort, while the average estimated effect was 0.078 NHI points per additional day of latency. These findings indicate that GA alone may not fully capture variation in neonatal outcomes of expectant management, and that additional clinical characteristics could help individualize management decisions.

Current guidelines recommend expectant management for PPROM before 34 weeks in the absence of contraindications.^4^ The greatest increase in predicted NHI with longer latency was observed among pregnancies with PPROM before 24 weeks’ gestation, consistent with prior reports of neonatal benefit near the limits of viability.^37,38^ In contrast, at later GA, predicted NHI was already high at short latency and increased only modestly with further latency, a pattern not captured by previous studies dichotomizing latency (e.g., <7 vs ≥7 days).^5–7^ Overall, these results indicate that the benefit of latency may not be adequately summarized by a single threshold or broad GA strata, as both its magnitude and pattern varied across finer GA subgroups.

Pregnancies in the highest-benefit quintile had a more favorable obstetric profile than those in the lowest, with lower rates of oligohydramnios, diabetes, hypertension, and multiple gestation, despite PPROM occurring at an earlier GA, a strong predictor of adverse neonatal outcomes (**Table 2**). This pattern suggests that pregnancies with earlier PPROM but otherwise favorable characteristics may have greater potential for improved neonatal outcomes with additional latency. Conversely, at later GA, predicted neonatal health already approached the upper limit of the NHI scale, leaving less measurable room for improvement with further latency (**Figure 1B**).

The identification of oligohydramnios and maternal age as key modifiers is biologically plausible: preserved amniotic fluid volume supports pulmonary and gastrointestinal maturation, and intrauterine immune defense, whereas reduced amniotic fluid volume has been associated with impaired lung development and adverse perinatal outcomes.^23,39^ Younger maternal age may also reflect greater placental reserve, consistent with reports of increased fetal vascular malperfusion from women aged 40 years or older,^40,41^ potentially limiting fetal perfusion as latency lengthens. Taken together, these findings suggest that longer latency may confer greater neonatal benefit in pregnancies with early PPROM occurring in a more preserved intrauterine environment, underscoring the importance of individualized risk stratification to refine expectant management beyond GA alone.

### Limitations

This study has several limitations. First, although our analyses were conducted within a causal inference framework, the observational design remains susceptible to residual confounding. Despite the robustness of the estimated APEs in sensitivity analyses, the registry lacked several potentially relevant obstetric factors, including transvaginal cervical length and cervical dilatation,^36,42,43^ maternal inflammatory markers (e.g., C-reactive protein, white blood cell count, and neutrophil-to-lymphocyte ratio),^44^ antenatal intrauterine infection,^45^ and initial fetal well-being assessments including non-stress test (NST) and biophysical profile (BPP).^46^ Clinical deterioration due to emerging infection, placental abruption, or fetal compromise can prompt earlier delivery and worsen neonatal outcomes,^3,4,36,47^ potentially favoring longer latency among clinically stable pregnancies. Secondly, the registry includes only neonates who survived to NICU admission, so neonates who died in the delivery room or before transfer to intensive care are absent. This may introduce selection bias, particular among the most immature or critically ill infants.^48^ Third, GA at PPROM was derived from clinical records and maternal reports, and therefore may be imprecise.^49^ The absence of a maternal identifier in the KNN registry also required use of a proxy, which may underestimate within-pregnancy correlation. Finally, because KNN includes only infants born before 32 weeks of gestation or weighing less than 1,500g, findings should be interpreted within this selected population and validated in independent populations.^50^

## Conclusions

The neonatal benefit associated with latency duration in PPROM was heterogeneous across patients, being greatest among those with earlier GA at PPROM, absent oligohydramnios, and younger maternal age. Considering these characteristics alongside GA may help individualize the expectant management of PPROM, though prospective validation is needed before clinical implementation.

## Data Availability

Due to ethical considerations regarding participant confidentiality and privacy, the Korean Neonatal Network (KNN) datasets generated and analyzed in this study are not publicly available. Requests for access may be directed to the corresponding author and will be reviewed in accordance with the KNN data-sharing policy.

## Author Contributions

SIS and YYJ designed the study; SIS acquired the study data; CS and JK performed the statistical analyses and generated the figures; CS, JK, SIS, and YYJ interpreted the results, drafted and reviewed the manuscript; YYJ acquired the funding. All authors reviewed and approved the final manuscript.

