## Supplementary Information for "Heterogeneous Associations of Latency Duration with Neonatal Outcomes After Preterm Prelabor Rupture of Membranes: A Nationwide Cohort Study"

### Supplementary Content

**Supplementary methods**

**Table S1.** Wald test *P* values for associations between covariates and neonatal outcomes in the full-cohort model from restricted cubic spline analysis

**Table S2.** Wald test *P* values for overall and nonlinear associations between latency period and neonatal outcomes in the subgroup-specific models from restricted cubic spline analysis

**Table S3.** Predicted Neonatal Health Index (NHI) at selected latency periods, derived from restricted cubic spline analysis

**Table S4**. Sensitivity analysis of the estimated average partial effect (APE) using the omitted variable bias (OVB) framework in causal machine learning implemented in EconML

**Table S5**. Estimated average partial effect (APE) and model calibration results of secondary outcomes

**Table S6**. Group average treatment effects (GATEs) for quartile-, quintile-, and decile-based stratifications of predicted individual treatment effects (ITEs)

**Table S7**. Pairwise contrasts in group average treatment effects (GATEs) comparing Q1 with higher quantile groups

**Table S8**. Variable importance

**Table S9**. Best linear projection (BLP) results

**Table S10**. Predicted Neonatal Health Index (NHI) at selected latency periods, derived from restricted cubic spline analysis with cluster-robust variance estimation (sensitivity analysis)

**Table S11**. Estimated average partial effects (APE) and model calibration results of the Neonatal Health Index (NHI) in the sensitivity analysis

**Table S12**. Group average treatment effects (GATEs) for quartile-, quintile-, and decile-based stratifications of predicted individual treatment effects (ITEs) in sensitivity analysis

**Table S13**. Pairwise contrasts in group average treatment effects (GATEs) comparing Q1 with higher quantile groups in sensitivity analysis

**Table S14**. Covariate comparison between the low (Q1) and high-benefit (Q5) group in GATE test in sensitivity analysis

**Table S15**. Variable importance obtained in the sensitivity analysis

**Table S16**. Best linear projection (BLP) results in the sensitivity analysis

**Figure S1.** Flowchart of study population selection

**Figure S2.** Group average treatment effects (GATEs) for quartile- and decile-based stratifications of predicted individual treatment effects (ITEs) of the NHI

**Figure S3.** Group average treatment effects (GATEs) for quintile-based stratifications of predicted individual treatment effects (ITEs) of morbidities and death used to construct the NHI

**Figure S4.** Variable importance of the secondary outcomes comprising the NHI

**Figure S5.** CATE interpreter trees derived from individual treatment effects (ITEs) estimated from the causal forest model for the NHI in EconML

**Figure S6.** Identification of variables contributing to treatment effect heterogeneity for the NHI in a sensitivity analysis restricting the sample to patients with latency period ≤ 30 days (N = 6,575)

**Abbreviations:** PPROM, Preterm prelabor rupture of membranes; GA, Gestational age; PH, Pulmonary Hypertension; BPD, Bronchopulmonary Dysplasia (moderate-severe); IVH, Intraventricular Hemorrhage (Grade 3-4); PVL, Periventricular Leukomalacia; SEPS, Sepsis; NEC, Necrotizing Enterocolitis (Stage ≥ 2); ROP, Retinopathy of Prematurity (Stage ≥ 3); NHI, Neonatal Health Index; KNN, Korean Neonatal Network; RCS, Restricted Cubic Spline; GRF, Generalized Random Forest; APE, Average Partial Effect; ITE, Individual Treatment Effect; CATE, Conditional Average Treatment Effect; GATE, Group Average Treatment Effect; BLP, Best Linear Predictor

#### Supplementary methods

##### Overview of the causal forest algorithm in continuous treatment setting and its implementation

We estimated the average treatment effect (ATE) and individual-level conditional average treatment effects (CATEs), hereafter referred to as ITEs individual treatment effects (ITEs), using causal forests within the generalized random forest (GRF) framework.

Three assumptions underlie this approach. First, ignorability, or the unconfoundedness assumption: conditional on the observed pre-treatment covariates, latency duration is independent of the potential neonatal outcomes. Although unconfoundedness cannot be verified empirically in observational data, we addressed it by including all clinically established determinants of both latency duration and neonatal morbidity available in the KNN registry and evaluating robustness to unobserved confounding through sensitivity analysis. Second, the positivity (overlap) assumption requires that each neonate has a non-negligible probability density of receiving its observed latency value conditional on the observed covariates. We estimated a generalized propensity score (GPS), f(latency duration | covariates), using a generalized linear model-based weighting approach (WeightIt R package, v1.7.0), which models the conditional density of the continuous exposure under a Gaussian residual assumption (**Figure, below**). In the full cohort (N = 7,057), the distribution of GPS-based stabilized weights indicated adequate covariate overlap for the large majority of neonates, with the low-density, high-weight observations concentrated in a small subgroup with markedly prolonged latency (GPS-weighted effective sample size [ESS]: 4,235, 60.0% of the nominal sample). Restricting the cohort to pregnancies with latency ≤ 30 days (n = 6,575) improved covariate overlap, increasing the effective sample size to 4,786 (72.8%), and supported the use of this restricted sample for sensitivity analysis. Third, the stable unit treatment value assumption (SUTVA): we assumed that the neonatal outcome of one neonate was not affected by the latency management of another.

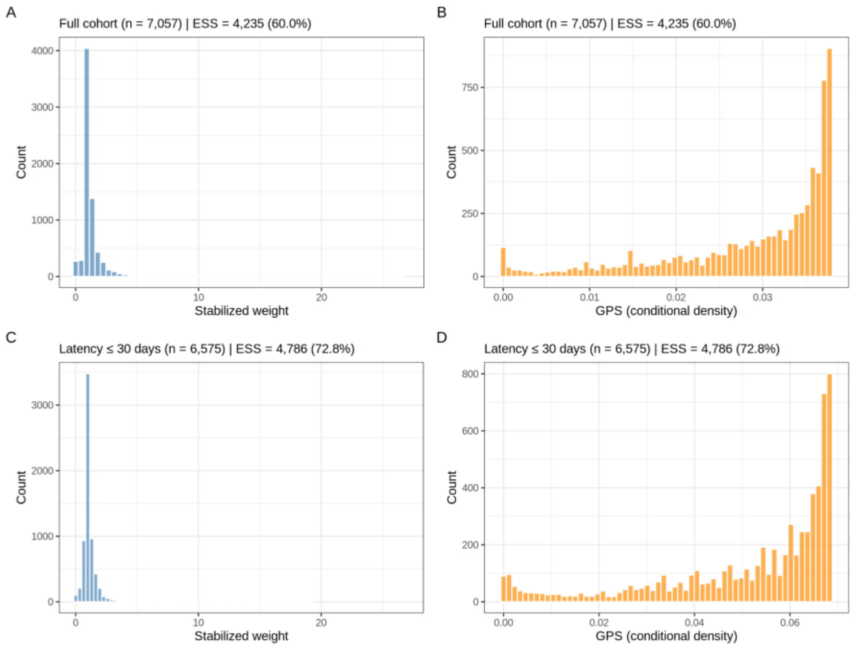

Causal forests employ 'honest' trees, in which each tree is grown on a subsample of the data that is randomly split into two disjoint parts: one used to build trees and the other to estimate treatment effects in leaf nodes. This property helps avoid overfitting by preventing the same sample from being used for both tree construction and effect estimation. During tree construction, splits are selected to maximize the difference in treatment effects between the two child nodes. Consequently, observations with similar covariate profiles are grouped together, and the treatment effect is assumed to be locally constant within each leaf node.

ITEs are estimated using out-of-bag (OOB) prediction. For a given sample $i$, it is passed through trees in which sample $i$ was not used at any stage of tree construction or estimation. For each eligible tree, samples that fall into the same leaf node as sample $i$ are identified. Aggregating across trees induces a set of forest-based adaptive weights $\alpha_j(x_i)$, defined by how frequently sample $j$ appears in the same leaf as sample $i$. These weights capture covariate similarity in a data-adaptive and nonlinear manner. Using these weights, the ITE for sample $i$ is estimated via a weighted residual-on-residual regression:

$$\tau(x) := \mathrm{lm}\big(Y_i - \hat{m}^{(-i)}(X_i) \ \sim \ W_i - \hat{e}^{(-i)}(X_i), \ \text{weights} = \alpha_i(x)\big)$$

where $Y_i$ denotes the outcome, $W_i$ the treatment, $\hat{m}(X) = \mathbb{E}[Y \mid X]$ the outcome nuisance model, and $\hat{e}(X) = \mathbb{E}[W \mid X]$ the treatment nuisance model (i.e., propensity score). The superscript $(-i)$ indicates cross-fitting, meaning that the nuisance predictions for sample $i$ are generated from models trained without using that sample.

As a result, treatment effect heterogeneity arises as $\tau(x)$ is modeled nonparametrically through recursive partitioning, enabling nonlinear interactions between covariates while remaining locally constant within each leaf node.

$$Y_i = \tau(x_i)W_i + f(x_i) + \epsilon_i$$

Here the treatment variable is continuous so the estimated CATE corresponds to a conditional partial effect, interpreted as a local slope

$$\tau(x) = \frac{\partial}{\partial w}\mathbb{E}[Y \mid X = x, W = w] = \frac{\mathrm{Cov}(Y, W \mid X = x)}{\mathrm{Var}(W \mid X = x)}.$$

The average treatment effect is therefore referred to as the average partial effect (APE). APE is obtained by averaging doubly robust (DR) scores rather than by simply averaging the ITEs.
Let

$$e(X) = \mathbb{E}[W \mid X], \quad v(X) = \mathrm{Var}(W \mid X), \quad m(W, X) = \mathbb{E}[Y \mid W, X]$$

In grf, the debiasing weights $v(X)$ are learned automatically by training auxiliary forests. We used the default settings debiasing.weights=NULL, num.trees.for.weights=500, which estimate the debiasing weights via 500 auxiliary forests. The DR score for a continuous treatment is given by

$$\psi_i = \frac{W_i - e(X_i)}{v(X_i)}\left(Y_i - m(W_i, X_i)\right) + \frac{\partial m(W_i, X_i)}{\partial W}.$$

The APE is then estimated as

$$\widehat{\mathrm{APE}} = \frac{1}{n}\sum_{i=1}^{n}\psi_i.$$

This estimator is doubly robust in the sense that it is consistent if either the treatment model $e(X)$, or the outcome model $m(W, X)$ is correctly specified. Under the unconfoundedness assumption, the estimated APE represents the average marginal effect of a one-unit increase in the continuous treatment (e.g., a one-day increase in latency duration) on the outcome. Unlike the *grf* package, *EconML* does not provide APE estimation based on averaging DR scores for continuous treatments.

Therefore it was obtained by directly averaging the ITEs estimated from CausalForestDML using const_marginal_effect().

Following prior methodological work1, we applied a seed-ensemble strategy to improve the stability of causal forest estimates in *grf*. This approach reduces sensitivity to random initialization and stabilizes calibration coefficients and ITE estimates without affecting bias. In practice, we trained forests using five independent random seeds with 2,000 trees each and merged them into a final ensemble forest comprising 10,000 trees.

As *grf* requires numeric inputs, categorical variables coded as 0 (no) and 1 (yes) were entered directly. One ordinal variable (small/appropriate/large for gestational age) was coded as 1,2,3 and entered directly because of its natural ordering. Categorical variables were one-hot encoded in *EconML*. Continuous variables were used on their original scales without standardization as tree-based methods are not sensitive to the scale of continuous predictors and to enhance interpretability.

**Model calibration test**

Model fit was assessed using a calibration test in *grf* evaluating whether the predicted APE and ITEs are consistent with the treatment-outcome relationship observed in the data.

$$Y_i - \hat{m}(X_i) = \beta_{\mathrm{ATE}}\,\hat{\tau}\left(W_i - \hat{e}(X_i)\right) + \beta_{\mathrm{ITE}}\left(\hat{\tau}_i - \hat{\tau}\right)\left(W_i - \hat{e}(X_i)\right) + \varepsilon$$

The coefficient $\beta_{\mathrm{ATE}}$ assesses calibration of the predicted APE, while $\beta_{\mathrm{ITE}}$ evaluates whether variation in predicted ITEs aligns with heterogeneity in the observed treatment effects. When these coefficients are close to 1 and statistically significant, this indicates that both the overall magnitude of the treatment effect and the pattern of treatment effect heterogeneity are well calibrated to the observed data.

**Cluster-based sensitivity analysis for within-pregnancy correlation**

Because neonates from multiple gestation share the same pregnancy-level exposure and may have correlated outcomes, we conducted cluster-based sensitivity analyses for both the restricted cubic spline (RCS) and causal forest analyses to account for within-pregnancy dependence that was not explicitly modeled in the primary analyses.[2,3] Because the registry did not include a maternal or pregnancy identifier, a proxy pregnancy identifier was constructed by grouping neonates with identical birth dates, maternal ages, parities, and gestational ages at delivery (week and day).

For the RCS analyses, models were repeated with the proxy pregnancy identifier as the clustering unit using cluster-robust variance estimation. Confidence intervals were obtained using cluster bootstrap resampling at the pregnancy level.[4]

For the *grf* analyses, the proxy pregnancy identifier was specified using the clustering variable ("clusters") so that pregnancy, rather than individual neonates, served as the sampling unit during forest construction. The "equalize.cluster.weights=TRUE" option was applied to ensure that each pregnancy contributed equally to forest estimation.

**Sensitivity analysis for unobserved confounding using the omitted variable bias framework in causal machine learning**

We applied the omitted variable bias sensitivity analysis implemented in *EconML*, which assesses the robustness of the estimated APE to potential unobserved confounding, to validate the APE estimate obtained from CausalForestDML. This framework evaluates whether an unobserved confounder is strong enough to overturn the estimated treatment effect.[5] The APE estimated from our fitted model (denoted as $\theta_s$) represents the treatment effect assuming that all relevant confounders have been measured. However, if unobserved confounders exist, the true treatment effect ($\theta$) may differ from $\theta_s$ due to omitted variable bias. Because the identity and strength of potential unobserved confounders are unknown, this method instead evaluates how strong such a confounder would need to be to invalidate the estimated effect. Confounding strength is characterized by two parameters: c_y and c_t, the strength of the unobserved confounder in explaining the outcome and the treatment, respectively. For example, c = 0.05 assumes that the unobserved confounder explains approximately 5% of the variation explained by the observed covariates. We conducted the analysis under moderate confounding assumptions (c_y = c_t = 0.05) and assumed worst-case alignment between treatment and outcome confounding ($\rho = 1$), which represents a conservative scenario where the unobserved confounder biases the treatment effect in the direction most adverse to the estimated effect.

Under these assumptions, we report the bias-adjusted APE ($\theta$), which represents the estimated treatment effect after accounting for potential unobserved confounding. The partially identified interval (PI) is the lower and upper bounds of $\theta$ under the specified confounding strength. We also report robustness values, which quantify the minimum strength of unobserved confounding required to invalidate the study conclusions. The robustness value for $\theta$ represents the confounding strength at which the bias-adjusted APE would become zero, whereas the robustness value for the 95% confidence interval (CI) represents the confounding strength at which the CI would include the null hypothesis (APE = 0). A larger robustness value indicates that stronger unobserved confounding would be required to overturn the estimated treatment effect.

**Table S1.** Wald test *P*-values for associations between covariates and neonatal outcomes in the full-cohort model from restricted cubic spline analysis. SGA and LGA were coded as dummy variables with AGA as the reference. We tested for nonlinearity only for the latency period, in which the *P*-values are presented in the last row.

| Covariate | NHI | IVH | PVL | ROP | BPD | PH | NEC | Sepsis | Death |
|---|---|---|---|---|---|---|---|---|---|
| Fetal sex | <.001 | .01 | .08 | .48 | <.001 | .11 | .049 | .14 | .003 |
| Multiple gestation | .07 | .04 | .95 | .09 | .78 | .89 | .48 | .76 | .91 |
| IVF conception | .19 | .65 | .54 | .95 | .86 | .72 | .53 | .21 | .94 |
| GDM / overt DM | .74 | .61 | .09 | .33 | .58 | .06 | .34 | .18 | .25 |
| Chronic HTN / PIH | .99 | .23 | .62 | .80 | .93 | .63 | .21 | .49 | .46 |
| Oligohydr amnios | <.001 | .08 | .87 | .41 | .03 | <.001 | .45 | .17 | .01 |
| Maternal age | .42 | .18 | .33 | .53 | .69 | .02 | .5 | .24 | .28 |
| Primiparit y | .14 | .77 | .50 | .13 | .32 | .04 | .9 | .93 | .65 |
| SGA | <.001 | .26 | .72 | .19 | <.001 | <.001 | .69 | .05 | <.001 |
| LGA | .2 | .09 | .35 | .47 | .19 | .82 | .27 | .21 | .91 |

| | | | | | | | | | |
|---|---|---|---|---|---|---|---|---|---|
| GA at PPROM (weeks) | <.001 | <.001 | <.001 | <.001 | <.001 | <.001 | <.001 | <.001 | <.001 |
| Latency Period | <.001 | <.001 | <.001 | <.001 | <.001 | <.001 | <.001 | <.001 | <.001 |
| Latency Period (Nonlinearity) | <.001 | <.001 | <.001 | .39 | .32 | .12 | .007 | <.001 | <.001 |

**Table S2.** Wald test *P* values for overall and nonlinear associations between latency period and neonatal outcomes in GA at PPROM subgroup-specific models from restricted cubic spline analyses.

| Outcome | <24 weeks | | 24-25+6/7 weeks | | 26-27+6/7 weeks | | >=28 weeks | |
|---|---|---|---|---|---|---|---|---|
| | overall *P* | nonlinearity *P* | overall *P* | nonlinearity *P* | overall *P* | nonlinearity *P* | overall *P* | nonlinearity *P* |
| NHI | < .001 | < .001 | < .001 | .004 | < .001 | < .001 | < .001 | .04 |
| IVH | < .001 | < .001 | < .001 | .56 | < .001 | .02 | < .001 | .001 |
| PVL | .01 | .47 | < .001 | .045 | < .001 | .17 | .005 | .05 |
| ROP | < .001 | .21 | < .001 | .07 | < .001 | .48 | .03 | .59 |
| BPD | < .001 | .30 | < .001 | .45 | < .001 | .92 | .002 | .97 |

| | | | | | | | | |
|---|---|---|---|---|---|---|---|---|
| PH | .15 | .30 | .002 | .49 | .02 | .16 | .1 | .77 |
| NEC | < .001 | .62 | < .001 | .004 | .002 | .24 | .6 | .78 |
| Sepsis | < .001 | .09 | < .001 | .33 | < .001 | .64 | < .001 | .23 |
| Death | < .001 | < .001 | < .001 | .15 | < .001 | .03 | .17 | .73 |

**Table S3.** Predicted Neonatal Health Index (NHI) at selected latency periods derived from restricted cubic spline analysis. Values are presented as predicted NHI (95% confidence interval); NHI ranges from 0 (worst) to 8 (best). A dash(—) indicates that the selected timepoint exceeded the 99th percentile of observed latency within that GA group.

| Latency period (days) | Full cohort model (GA-adjusted, n=6,872) | Subgroup-specific models | | | |
|---|---|---|---|---|---|
| | | <24 weeks (n=1,270) | 24–25+6 weeks (n=1,238) | 26–27+6 weeks (n=1,423) | ≥28 weeks (n=2,941) |
| | Predicted NHI (95% CI) | Predicted NHI (95% CI) | Predicted NHI (95% CI) | Predicted NHI (95% CI) | Predicted NHI (95% CI) |
| 0 | 5.19 [5.09-5.28] | 1.36 [1.10-1.66] | 3.81 [3.52-4.08] | 5.86 [5.67-6.06] | 7.28 [7.21-7.36] |
| 1 | 5.44 [5.36-5.53] | 1.60 [1.36-1.85] | 4.08 [3.88-4.27] | 6.24 [6.10-6.38] | 7.37 [7.30-7.42] |
| 2 | 5.66 [5.58-5.75] | 1.87 [1.63-2.10] | 4.33 [4.10-4.56] | 6.52 [6.34-6.68] | 7.45 [7.37-7.50] |
| 3 | 5.77 [5.69-5.86] | 2.14 [1.89-2.38] | 4.58 [4.30-4.85] | 6.68 [6.50-6.85] | 7.50 [7.42-7.56] |

| | | | | | |
|---|---|---|---|---|---|
| 4 | 5.90 [5.81-5.98] | 2.40 [2.12-2.67] | 4.81 [4.50-5.12] | 6.78 [6.61-6.92] | 7.54 [7.47-7.61] |
| 5 | 6.00 [5.91-6.08] | 2.63 [2.33-2.94] | 5.03 [4.71-5.34] | 6.81 [6.66-6.94] | 7.57 [7.50-7.64] |
| 6 | 6.07 [5.99-6.15] | 2.84 [2.51-3.17] | 5.23 [4.92-5.52] | 6.83 [6.68-6.96] | 7.60 [7.53-7.66] |
| 7 | 6.12 [6.04-6.19] | 3.02 [2.69-3.37] | 5.41 [5.14-5.68] | 6.86 [6.69-7.00] | 7.61 [7.55-7.68] |
| 14 | 6.51 [6.44-6.59] | 3.69 [3.44-4.01] | 6.30 [6.02-6.57] | 7.17 [7.01-7.30] | 7.70 [7.56-7.83] |
| 21 | 6.82 [6.74-6.89] | 3.98 [3.65-4.35] | 6.74 [6.54-6.95] | 7.50 [7.36-7.64] | — |
| 28 | 7.06 [7.00-7.13] | 4.30 [3.99-4.63] | 7.04 [6.77-7.26] | 7.71 [7.55-7.84] | — |
| 35 | 7.28 [7.23-7.33] | 4.61 [4.32-4.91] | 7.26 [6.88-7.54] | — | — |
| 42 | 7.44 [7.40-7.49] | 4.91 [4.56-5.25] | — | — | — |
| 49 | 7.57 [7.53-7.61] | 5.15 [4.78-5.51] | — | — | — |
| 56 | 7.68 [7.64-7.71] | 5.38 [5.02-5.72] | — | — | — |
| 63 | 7.76 [7.72-7.79] | 5.57 [5.23-5.92] | — | — | — |
| 70 | 7.82 [7.79-7.85] | 5.74 [5.34-6.09] | — | — | — |
| 77 | — | 5.90 [5.44-6.28] | — | — | — |
| 84 | — | 6.07 [5.49-6.52] | — | — | — |
| 91 | — | 6.21 [5.56-6.74] | — | — | — |

| 97 | — | 6.33 [5.59-6.90] | — | — | — |
|---|---|---|---|---|---|

**Table S4**. Sensitivity analysis of the estimated average partial effect (APE) using the omitted variable bias (OVB) framework in causal machine learning implemented in EconML.

| Bias-adjusted APE (θ) | Partially Identified Interval (PI) | 95% Confidence Interval (CI) | Robustness Value of bias-adjusted APE | Robustness Value of 95% CI |
|---|---|---|---|---|
| 0.116 | [0.097, 0.135] | [0.088, 0.144] | 0.269 | 0.249 |

**Table S5**. Estimated average partial effect (APE) and model calibration results of secondary outcomes.

| | grf | | | | | | EconML | |
|---|---|---|---|---|---|---|---|---|
| Outcome | APE | p | βATE | p | βITE | p | APE | p |
| PH | 0.0033 | <.001 | 1.85 | <.001 | -179.48 | >.99 | 0.0042 | <.001 |
| BPD | 0.0113 | <.001 | 1.07 | <.001 | 2.08 | .006 | 0.0116 | <.001 |
| IVH | 0.0055 | <.001 | 1.01 | <.001 | 1.85 | <.001 | 0.0059 | <.001 |
| PVL | 0.0044 | <.001 | 0.73 | <.001 | -9.92 | .99 | 0.0032 | .007 |
| Sepsis | 0.0085 | <.001 | 1.09 | <.001 | 1.82 | .046 | 0.0078 | <.001 |
| NEC | 0.0017 | .02 | 0.98 | <.001 | -6.00 | .96 | 0.0032 | <.001 |
| ROP | 0.0069 | <.001 | 0.82 | <.001 | 1.82 | <.001 | 0.0074 | .04 |

| | | | | | | | | |
|---|---|---|---|---|---|---|---|---|
| Death | 0.0066 | <.001 | 0.85 | <.001 | 1.26 | <.001 | 0.0071 | <.001 |

**Table S6.** Group average treatment effects (GATEs) for quartile-, quintile-, and decile-based stratifications of predicted individual treatment effects (ITEs). P-values test the null hypothesis that the GATE equals zero using two-sided Wald tests. n.s., not significant; *** p < .001; ** p < .01; * p < .05.

| Outcome | Quantile type | Quantile group | CI_low | CI_high | GATE_est | GATE_se | GATE_p | Significance |
|---|---|---|---|---|---|---|---|---|
| PH | quartile | Q1 | 0.0038 | 0.0039 | 0.0049 | 0.0009 | <.001 | *** |
| PH | quartile | Q2 | 0.0039 | 0.0040 | 0.0070 | 0.0009 | <.001 | *** |
| PH | quartile | Q3 | 0.0040 | 0.0040 | 0.0025 | 0.0010 | .02 | * |
| PH | quartile | Q4 | 0.0040 | 0.0041 | -0.0014 | 0.0010 | .18 | n.s. |
| PH | quintile | Q1 | 0.0038 | 0.0039 | 0.0053 | 0.0011 | <.001 | *** |
| PH | quintile | Q2 | 0.0039 | 0.0039 | 0.0073 | 0.0011 | <.001 | *** |
| PH | quintile | Q3 | 0.0039 | 0.0040 | 0.0034 | 0.0010 | <.001 | *** |
| PH | quintile | Q4 | 0.0040 | 0.0040 | 0.0037 | 0.0011 | <.001 | *** |
| PH | quintile | Q5 | 0.0040 | 0.0041 | -0.0035 | 0.0012 | .003 | ** |
| PH | decile | Q1 | 0.0038 | 0.0039 | 0.0056 | 0.0017 | <.001 | *** |
| PH | decile | Q2 | 0.0039 | 0.0039 | 0.0051 | 0.0013 | <.001 | *** |

| | | | | | | | | |
|---|---|---|---|---|---|---|---|---|
| PH | decile | Q3 | 0.0039 | 0.0039 | 0.0076 | 0.0013 | <.001 | *** |
| PH | decile | Q4 | 0.0039 | 0.0039 | 0.0070 | 0.0018 | <.001 | *** |
| PH | decile | Q5 | 0.0039 | 0.0040 | 0.0044 | 0.0010 | <.001 | *** |
| PH | decile | Q6 | 0.0040 | 0.0040 | 0.0024 | 0.0017 | .16 | n.s. |
| PH | decile | Q7 | 0.0040 | 0.0040 | 0.0034 | 0.0017 | .04 | * |
| PH | decile | Q8 | 0.0040 | 0.0040 | 0.0040 | 0.0013 | .002 | ** |
| PH | decile | Q9 | 0.0040 | 0.0040 | 0.0020 | 0.0016 | .21 | n.s. |
| PH | decile | Q10 | 0.0040 | 0.0041 | -0.0090 | 0.0018 | <.001 | *** |
| BPD | quartile | Q1 | 0.0095 | 0.0110 | 0.0091 | 0.0017 | <.001 | *** |
| BPD | quartile | Q2 | 0.0110 | 0.0117 | 0.0088 | 0.0025 | <.001 | *** |
| BPD | quartile | Q3 | 0.0117 | 0.0124 | 0.0108 | 0.0025 | <.001 | *** |
| BPD | quartile | Q4 | 0.0124 | 0.0139 | 0.0164 | 0.0025 | <.001 | *** |
| BPD | quintile | Q1 | 0.0095 | 0.0109 | 0.0087 | 0.0018 | <.001 | *** |
| BPD | quintile | Q2 | 0.0109 | 0.0115 | 0.0085 | 0.0026 | .001 | ** |
| BPD | quintile | Q3 | 0.0115 | 0.0119 | 0.0118 | 0.0025 | <.001 | *** |
| BPD | quintile | Q4 | 0.0119 | 0.0126 | 0.0118 | 0.0029 | <.001 | *** |

| | | | | | | | | |
|---|---|---|---|---|---|---|---|---|
| BPD | quintile | Q5 | 0.0126 | 0.0139 | 0.0156 | 0.0030 | <.001 | *** |
| BPD | decile | Q1 | 0.0095 | 0.0106 | 0.0070 | 0.0022 | .001 | ** |
| BPD | decile | Q2 | 0.0106 | 0.0109 | 0.0104 | 0.0028 | <.001 | *** |
| BPD | decile | Q3 | 0.0109 | 0.0112 | 0.0107 | 0.0033 | .001 | ** |
| BPD | decile | Q4 | 0.0112 | 0.0115 | 0.0062 | 0.0040 | .12 | n.s. |
| BPD | decile | Q5 | 0.0115 | 0.0117 | 0.0105 | 0.0042 | .01 | * |
| BPD | decile | Q6 | 0.0117 | 0.0119 | 0.0132 | 0.0028 | <.001 | *** |
| BPD | decile | Q7 | 0.0119 | 0.0122 | 0.0070 | 0.0051 | .17 | n.s. |
| BPD | decile | Q8 | 0.0122 | 0.0126 | 0.0165 | 0.0030 | <.001 | *** |
| BPD | decile | Q9 | 0.0126 | 0.0129 | 0.0207 | 0.0035 | <.001 | *** |
| BPD | decile | Q10 | 0.0129 | 0.0139 | 0.0105 | 0.0048 | .03 | * |
| IVH | quartile | Q1 | 0.0047 | 0.0063 | 0.0046 | 0.0009 | <.001 | *** |
| IVH | quartile | Q2 | 0.0063 | 0.0067 | 0.0031 | 0.0009 | <.001 | *** |
| IVH | quartile | Q3 | 0.0067 | 0.0072 | 0.0057 | 0.0008 | <.001 | *** |
| IVH | quartile | Q4 | 0.0072 | 0.0084 | 0.0084 | 0.0008 | <.001 | *** |
| IVH | quintile | Q1 | 0.0047 | 0.0061 | 0.0043 | 0.0010 | <.001 | *** |

| | | | | | | | | |
|---|---|---|---|---|---|---|---|---|
| IVH | quintile | Q2 | 0.0061 | 0.00 66 | 0.00 36 | 0.00 10 | <.001 | *** |
| IVH | quintile | Q3 | 0.0066 | 0.00 68 | 0.00 38 | 0.00 09 | <.001 | *** |
| IVH | quintile | Q4 | 0.0068 | 0.00 73 | 0.00 74 | 0.00 10 | <.001 | *** |
| IVH | quintile | Q5 | 0.0073 | 0.00 84 | 0.00 83 | 0.00 09 | <.001 | *** |
| IVH | decile | Q1 | 0.0047 | 0.00 54 | 0.00 40 | 0.00 17 | .02 | * |
| IVH | decile | Q2 | 0.0054 | 0.00 61 | 0.00 47 | 0.00 11 | <.001 | *** |
| IVH | decile | Q3 | 0.0061 | 0.00 64 | 0.00 59 | 0.00 13 | <.001 | *** |
| IVH | decile | Q4 | 0.0064 | 0.00 66 | 0.00 13 | 0.00 14 | .37 | n.s. |
| IVH | decile | Q5 | 0.0066 | 0.00 67 | 0.00 37 | 0.00 14 | .007 | ** |
| IVH | decile | Q6 | 0.0067 | 0.00 68 | 0.00 39 | 0.00 13 | .002 | ** |
| IVH | decile | Q7 | 0.0068 | 0.00 70 | 0.00 66 | 0.00 15 | <.001 | *** |
| IVH | decile | Q8 | 0.0070 | 0.00 73 | 0.00 82 | 0.00 14 | <.001 | *** |
| IVH | decile | Q9 | 0.0073 | 0.00 76 | 0.00 77 | 0.00 13 | <.001 | *** |
| IVH | decile | Q10 | 0.0076 | 0.00 84 | 0.00 89 | 0.00 12 | <.001 | *** |
| PVL | quartile | Q1 | 0.0027 | 0.00 30 | 0.00 42 | 0.00 06 | <.001 | *** |
| PVL | quartile | Q2 | 0.0030 | 0.00 31 | 0.00 55 | 0.00 11 | <.001 | *** |

| | | | | | | | | |
|---|---|---|---|---|---|---|---|---|
| PVL | quartile | Q3 | 0.0031 | 0.0031 | 0.0051 | 0.0012 | <.001 | *** |
| PVL | quartile | Q4 | 0.0031 | 0.0033 | 0.0027 | 0.0015 | .08 | n.s. |
| PVL | quintile | Q1 | 0.0027 | 0.0030 | 0.0048 | 0.0006 | <.001 | *** |
| PVL | quintile | Q2 | 0.0030 | 0.0030 | 0.0043 | 0.0011 | <.001 | *** |
| PVL | quintile | Q3 | 0.0030 | 0.0031 | 0.0052 | 0.0013 | <.001 | *** |
| PVL | quintile | Q4 | 0.0031 | 0.0032 | 0.0059 | 0.0014 | <.001 | *** |
| PVL | quintile | Q5 | 0.0032 | 0.0033 | 0.0018 | 0.0018 | .32 | n.s. |
| PVL | decile | Q1 | 0.0027 | 0.0029 | 0.0055 | 0.0008 | <.001 | *** |
| PVL | decile | Q2 | 0.0029 | 0.0030 | 0.0041 | 0.0008 | <.001 | *** |
| PVL | decile | Q3 | 0.0030 | 0.0030 | 0.0031 | 0.0012 | .010 | ** |
| PVL | decile | Q4 | 0.0030 | 0.0030 | 0.0056 | 0.0019 | .003 | ** |
| PVL | decile | Q5 | 0.0030 | 0.0031 | 0.0061 | 0.0018 | <.001 | *** |
| PVL | decile | Q6 | 0.0031 | 0.0031 | 0.0043 | 0.0019 | .03 | * |
| PVL | decile | Q7 | 0.0031 | 0.0031 | 0.0071 | 0.0020 | <.001 | *** |
| PVL | decile | Q8 | 0.0031 | 0.0032 | 0.0047 | 0.0020 | .02 | * |
| PVL | decile | Q9 | 0.0032 | 0.0032 | -0.0012 | 0.0028 | .67 | n.s. |

| | | | | | | | | |
|---|---|---|---|---|---|---|---|---|
| PVL | decile | Q10 | 0.0032 | 0.00 33 | 0.00 47 | 0.00 22 | .03 | * |
| SEPS | quartile | Q1 | 0.0056 | 0.00 66 | 0.00 75 | 0.00 09 | <.001 | *** |
| SEPS | quartile | Q2 | 0.0066 | 0.00 69 | 0.01 09 | 0.00 13 | <.001 | *** |
| SEPS | quartile | Q3 | 0.0069 | 0.00 72 | 0.00 94 | 0.00 15 | <.001 | *** |
| SEPS | quartile | Q4 | 0.0072 | 0.00 76 | 0.00 63 | 0.00 17 | <.001 | *** |
| SEPS | quintile | Q1 | 0.0056 | 0.00 65 | 0.00 76 | 0.00 09 | <.001 | *** |
| SEPS | quintile | Q2 | 0.0065 | 0.00 68 | 0.01 05 | 0.00 14 | <.001 | *** |
| SEPS | quintile | Q3 | 0.0068 | 0.00 70 | 0.01 06 | 0.00 17 | <.001 | *** |
| SEPS | quintile | Q4 | 0.0070 | 0.00 72 | 0.00 87 | 0.00 17 | <.001 | *** |
| SEPS | quintile | Q5 | 0.0072 | 0.00 76 | 0.00 52 | 0.00 18 | .003 | ** |
| SEPS | decile | Q1 | 0.0056 | 0.00 62 | 0.00 63 | 0.00 11 | <.001 | *** |
| SEPS | decile | Q2 | 0.0062 | 0.00 65 | 0.00 89 | 0.00 13 | <.001 | *** |
| SEPS | decile | Q3 | 0.0065 | 0.00 67 | 0.01 01 | 0.00 19 | <.001 | *** |
| SEPS | decile | Q4 | 0.0067 | 0.00 68 | 0.01 08 | 0.00 22 | <.001 | *** |
| SEPS | decile | Q5 | 0.0068 | 0.00 69 | 0.00 98 | 0.00 22 | <.001 | *** |
| SEPS | decile | Q6 | 0.0069 | 0.00 70 | 0.01 15 | 0.00 27 | <.001 | *** |

| | | | | | | | | |
|---|---|---|---|---|---|---|---|---|
| SEPS | decile | Q7 | 0.0070 | 0.0071 | 0.0066 | 0.0020 | .001 | ** |
| SEPS | decile | Q8 | 0.0071 | 0.0072 | 0.0107 | 0.0028 | <.001 | *** |
| SEPS | decile | Q9 | 0.0072 | 0.0073 | 0.0098 | 0.0023 | <.001 | *** |
| SEPS | decile | Q10 | 0.0073 | 0.0076 | 0.0006 | 0.0027 | .83 | n.s. |
| NEC | quartile | Q1 | 0.0027 | 0.0029 | 0.0013 | 0.0026 | .62 | n.s. |
| NEC | quartile | Q2 | 0.0029 | 0.0031 | 0.0013 | 0.0007 | .051 | n.s. |
| NEC | quartile | Q3 | 0.0031 | 0.0032 | 0.0028 | 0.0010 | .003 | ** |
| NEC | quartile | Q4 | 0.0032 | 0.0034 | 0.0015 | 0.0010 | .16 | n.s. |
| NEC | quintile | Q1 | 0.0027 | 0.0029 | 0.0014 | 0.0032 | .65 | n.s. |
| NEC | quintile | Q2 | 0.0029 | 0.0030 | 0.0011 | 0.0007 | .14 | n.s. |
| NEC | quintile | Q3 | 0.0030 | 0.0031 | 0.0025 | 0.0008 | .002 | ** |
| NEC | quintile | Q4 | 0.0031 | 0.0032 | 0.0017 | 0.0014 | .21 | n.s. |
| NEC | quintile | Q5 | 0.0032 | 0.0034 | 0.0018 | 0.0010 | .06 | n.s. |
| NEC | decile | Q1 | 0.0027 | 0.0028 | 0.0050 | 0.0017 | .003 | ** |
| NEC | decile | Q2 | 0.0028 | 0.0029 | -0.0022 | 0.0061 | .72 | n.s. |
| NEC | decile | Q3 | 0.0029 | 0.0030 | 0.0016 | 0.0012 | .18 | n.s. |

| | | | | | | | | |
|---|---|---|---|---|---|---|---|---|
| NEC | decile | Q4 | 0.0030 | 0.0030 | 0.0007 | 0.0009 | .48 | n.s. |
| NEC | decile | Q5 | 0.0030 | 0.0031 | 0.0014 | 0.0012 | .26 | n.s. |
| NEC | decile | Q6 | 0.0031 | 0.0031 | 0.0037 | 0.0011 | .001 | ** |
| NEC | decile | Q7 | 0.0031 | 0.0031 | 0.0018 | 0.0018 | .33 | n.s. |
| NEC | decile | Q8 | 0.0031 | 0.0032 | 0.0017 | 0.0021 | .41 | n.s. |
| NEC | decile | Q9 | 0.0032 | 0.0033 | 0.0006 | 0.0014 | .65 | n.s. |
| NEC | decile | Q10 | 0.0033 | 0.0034 | 0.0030 | 0.0013 | .02 | * |
| ROP | quartile | Q1 | 0.0063 | 0.0073 | 0.0031 | 0.0009 | <.001 | *** |
| ROP | quartile | Q2 | 0.0073 | 0.0084 | 0.0034 | 0.0012 | .004 | ** |
| ROP | quartile | Q3 | 0.0084 | 0.0113 | 0.0086 | 0.0008 | <.001 | *** |
| ROP | quartile | Q4 | 0.0113 | 0.0143 | 0.0123 | 0.0011 | <.001 | *** |
| ROP | quintile | Q1 | 0.0063 | 0.0072 | 0.0029 | 0.0011 | .005 | ** |
| ROP | quintile | Q2 | 0.0072 | 0.0080 | 0.0037 | 0.0011 | <.001 | *** |
| ROP | quintile | Q3 | 0.0080 | 0.0108 | 0.0030 | 0.0013 | .02 | * |
| ROP | quintile | Q4 | 0.0108 | 0.0116 | 0.0112 | 0.0009 | <.001 | *** |
| ROP | quintile | Q5 | 0.0116 | 0.0143 | 0.0135 | 0.0012 | <.001 | *** |

| | | | | | | | | |
|---|---|---|---|---|---|---|---|---|
| ROP | decile | Q1 | 0.0063 | 0.0069 | 0.0035 | 0.0015 | .02 | * |
| ROP | decile | Q2 | 0.0069 | 0.0072 | 0.0024 | 0.0015 | .09 | n.s. |
| ROP | decile | Q3 | 0.0072 | 0.0075 | 0.0027 | 0.0013 | .03 | * |
| ROP | decile | Q4 | 0.0075 | 0.0080 | 0.0048 | 0.0018 | .010 | ** |
| ROP | decile | Q5 | 0.0080 | 0.0084 | 0.0028 | 0.0021 | .19 | n.s. |
| ROP | decile | Q6 | 0.0084 | 0.0108 | 0.0032 | 0.0015 | .03 | * |
| ROP | decile | Q7 | 0.0108 | 0.0111 | 0.0124 | 0.0011 | <.001 | *** |
| ROP | decile | Q8 | 0.0111 | 0.0116 | 0.0099 | 0.0014 | <.001 | *** |
| ROP | decile | Q9 | 0.0116 | 0.0131 | 0.0115 | 0.0019 | <.001 | *** |
| ROP | decile | Q10 | 0.0131 | 0.0143 | 0.0155 | 0.0015 | <.001 | *** |
| Death | quartile | Q1 | 0.0039 | 0.0053 | 0.0036 | 0.0010 | <.001 | *** |
| Death | quartile | Q2 | 0.0053 | 0.0063 | 0.0027 | 0.0010 | .008 | ** |
| Death | quartile | Q3 | 0.0063 | 0.0099 | 0.0064 | 0.0009 | <.001 | *** |
| Death | quartile | Q4 | 0.0099 | 0.0157 | 0.0136 | 0.0007 | <.001 | *** |
| Death | quintile | Q1 | 0.0039 | 0.0052 | 0.0029 | 0.0010 | .005 | ** |
| Death | quintile | Q2 | 0.0052 | 0.0059 | 0.0030 | 0.0012 | .02 | * |

| | | | | | | | | |
|---|---|---|---|---|---|---|---|---|
| Death | quintile | Q3 | 0.0059 | 0.00 67 | 0.00 41 | 0.00 09 | <.001 | *** |
| Death | quintile | Q4 | 0.0067 | 0.01 04 | 0.00 85 | 0.00 11 | <.001 | *** |
| Death | quintile | Q5 | 0.0104 | 0.01 57 | 0.01 43 | 0.00 08 | <.001 | *** |
| Death | decile | Q1 | 0.0039 | 0.00 48 | 0.00 21 | 0.00 15 | .18 | n.s. |
| Death | decile | Q2 | 0.0048 | 0.00 52 | 0.00 38 | 0.00 14 | .007 | ** |
| Death | decile | Q3 | 0.0052 | 0.00 55 | 0.00 24 | 0.00 18 | .17 | n.s. |
| Death | decile | Q4 | 0.0055 | 0.00 59 | 0.00 35 | 0.00 17 | .04 | * |
| Death | decile | Q5 | 0.0059 | 0.00 63 | 0.00 40 | 0.00 14 | .005 | ** |
| Death | decile | Q6 | 0.0063 | 0.00 67 | 0.00 43 | 0.00 12 | <.001 | *** |
| Death | decile | Q7 | 0.0067 | 0.00 89 | 0.00 51 | 0.00 17 | .002 | ** |
| Death | decile | Q8 | 0.0089 | 0.01 04 | 0.01 19 | 0.00 14 | <.001 | *** |
| Death | decile | Q9 | 0.0104 | 0.01 45 | 0.01 09 | 0.00 12 | <.001 | *** |
| Death | decile | Q10 | 0.0145 | 0.01 57 | 0.01 78 | 0.00 11 | <.001 | *** |
| NHI | quartile | Q1 | 0.0650 | 0.08 31 | 0.04 12 | 0.01 00 | <.001 | *** |
| NHI | quartile | Q2 | 0.0831 | 0.09 08 | 0.05 85 | 0.01 18 | <.001 | *** |
| NHI | quartile | Q3 | 0.0908 | 0.10 69 | 0.07 73 | 0.00 69 | <.001 | *** |

| | | | | | | | | |
|---|---|---|---|---|---|---|---|---|
| NHI | quartile | Q4 | 0.1069 | 0.1438 | 0.1335 | 0.0055 | <.001 | *** |
| NHI | quintile | Q1 | 0.0650 | 0.0818 | 0.0406 | 0.0115 | <.001 | *** |
| NHI | quintile | Q2 | 0.0818 | 0.0876 | 0.0584 | 0.0127 | <.001 | *** |
| NHI | quintile | Q3 | 0.0876 | 0.0953 | 0.0562 | 0.0105 | <.001 | *** |
| NHI | quintile | Q4 | 0.0953 | 0.1117 | 0.0993 | 0.0076 | <.001 | *** |
| NHI | quintile | Q5 | 0.1117 | 0.1438 | 0.1335 | 0.0059 | <.001 | *** |
| NHI | decile | Q1 | 0.0650 | 0.0777 | 0.0316 | 0.0203 | .12 | n.s. |
| NHI | decile | Q2 | 0.0777 | 0.0818 | 0.0497 | 0.0107 | <.001 | *** |
| NHI | decile | Q3 | 0.0818 | 0.0847 | 0.0450 | 0.0198 | .02 | * |
| NHI | decile | Q4 | 0.0847 | 0.0876 | 0.0718 | 0.0161 | <.001 | *** |
| NHI | decile | Q5 | 0.0876 | 0.0908 | 0.0511 | 0.0181 | .005 | ** |
| NHI | decile | Q6 | 0.0908 | 0.0953 | 0.0614 | 0.0106 | <.001 | *** |
| NHI | decile | Q7 | 0.0953 | 0.1021 | 0.0844 | 0.0122 | <.001 | *** |
| NHI | decile | Q8 | 0.1021 | 0.1117 | 0.1143 | 0.0089 | <.001 | *** |
| NHI | decile | Q9 | 0.1117 | 0.1213 | 0.1191 | 0.0084 | <.001 | *** |
| NHI | decile | Q10 | 0.1213 | 0.1438 | 0.1478 | 0.0084 | <.001 | *** |

**Table S7.** Pairwise contrasts in group average treatment effects (GATEs) comparing Q1 with higher quantile groups. For each outcome, contrasts were calculated as the difference in estimated GATE between each subgroup and Q1 using Wald tests. n.s., not significant; *** p < .001; ** p < .01; * p < .05.

| Outcome | Quantile type | contrast | diff | se | z | p | Significance |
|---|---|---|---|---|---|---|---|
| PH | quartile | Q2 - Q1 | 0.002 | 0.001 | 1.66 | .10 | n.s. |
| PH | quartile | Q3 - Q1 | -0.002 | 0.001 | -1.71 | .09 | n.s. |
| PH | quartile | Q4 - Q1 | -0.006 | 0.001 | -4.50 | <.001 | *** |
| PH | quintile | Q2 - Q1 | 0.002 | 0.002 | 1.28 | .20 | n.s. |
| PH | quintile | Q3 - Q1 | -0.002 | 0.001 | -1.32 | .19 | n.s. |
| PH | quintile | Q4 - Q1 | -0.002 | 0.002 | -1.07 | .28 | n.s. |
| PH | quintile | Q5 - Q1 | -0.009 | 0.002 | -5.52 | <.001 | *** |
| PH | decile | Q2 - Q1 | 0.000 | 0.002 | -0.21 | .83 | n.s. |
| PH | decile | Q3 - Q1 | 0.002 | 0.002 | 0.96 | .34 | n.s. |
| PH | decile | Q4 - Q1 | 0.001 | 0.002 | 0.59 | .56 | n.s. |
| PH | decile | Q5 - Q1 | -0.001 | 0.002 | -0.57 | .57 | n.s. |
| PH | decile | Q6 - Q1 | -0.003 | 0.002 | -1.33 | .18 | n.s. |
| PH | decile | Q7 - Q1 | -0.002 | 0.002 | -0.89 | .37 | n.s. |

| | | | | | | | |
|---|---|---|---|---|---|---|---|
| PH | decile | Q8 - Q1 | -0.002 | 0.002 | -0.74 | .46 | n.s. |
| PH | decile | Q9 - Q1 | -0.004 | 0.002 | -1.55 | .12 | n.s. |
| PH | decile | Q10 - Q1 | -0.015 | 0.002 | -6.00 | <.001 | *** |
| BPD | quartile | Q2 - Q1 | 0.000 | 0.003 | -0.08 | .94 | n.s. |
| BPD | quartile | Q3 - Q1 | 0.002 | 0.003 | 0.57 | .57 | n.s. |
| BPD | quartile | Q4 - Q1 | 0.007 | 0.003 | 2.38 | .02 | * |
| BPD | quintile | Q2 - Q1 | 0.000 | 0.003 | -0.08 | .93 | n.s. |
| BPD | quintile | Q3 - Q1 | 0.003 | 0.003 | 1.00 | .32 | n.s. |
| BPD | quintile | Q4 - Q1 | 0.003 | 0.003 | 0.88 | .38 | n.s. |
| BPD | quintile | Q5 - Q1 | 0.007 | 0.003 | 1.98 | .048 | * |
| BPD | decile | Q2 - Q1 | 0.003 | 0.004 | 0.95 | .34 | n.s. |
| BPD | decile | Q3 - Q1 | 0.004 | 0.004 | 0.94 | .35 | n.s. |
| BPD | decile | Q4 - Q1 | -0.001 | 0.005 | -0.17 | .86 | n.s. |
| BPD | decile | Q5 - Q1 | 0.003 | 0.005 | 0.73 | .47 | n.s. |
| BPD | decile | Q6 - Q1 | 0.006 | 0.004 | 1.72 | .09 | n.s. |
| BPD | decile | Q7 - Q1 | 0.000 | 0.006 | 0.00 | 1.00 | n.s. |

| | | | | | | | |
|---|---|---|---|---|---|---|---|
| BPD | decile | Q8 - Q1 | 0.009 | 0.004 | 2.54 | .01 | * |
| BPD | decile | Q9 - Q1 | 0.014 | 0.004 | 3.30 | .001 | ** |
| BPD | decile | Q10 - Q1 | 0.003 | 0.005 | 0.65 | .52 | n.s. |
| IVH | quartile | Q2 - Q1 | -0.002 | 0.001 | -1.23 | .22 | n.s. |
| IVH | quartile | Q3 - Q1 | 0.001 | 0.001 | 0.90 | .37 | n.s. |
| IVH | quartile | Q4 - Q1 | 0.004 | 0.001 | 3.11 | .002 | ** |
| IVH | quintile | Q2 - Q1 | -0.001 | 0.001 | -0.54 | .59 | n.s. |
| IVH | quintile | Q3 - Q1 | 0.000 | 0.001 | -0.36 | .72 | n.s. |
| IVH | quintile | Q4 - Q1 | 0.003 | 0.001 | 2.17 | .03 | * |
| IVH | quintile | Q5 - Q1 | 0.004 | 0.001 | 2.98 | .003 | ** |
| IVH | decile | Q2 - Q1 | 0.001 | 0.002 | 0.34 | .73 | n.s. |
| IVH | decile | Q3 - Q1 | 0.002 | 0.002 | 0.91 | .36 | n.s. |
| IVH | decile | Q4 - Q1 | -0.003 | 0.002 | -1.23 | .22 | n.s. |
| IVH | decile | Q5 - Q1 | 0.000 | 0.002 | -0.12 | .91 | n.s. |
| IVH | decile | Q6 - Q1 | 0.000 | 0.002 | -0.02 | .98 | n.s. |
| IVH | decile | Q7 - Q1 | 0.003 | 0.002 | 1.17 | .24 | n.s. |

| | | | | | | | |
|---|---|---|---|---|---|---|---|
| IVH | decile | Q8 - Q1 | 0.004 | 0.002 | 1.95 | .051 | n.s. |
| IVH | decile | Q9 - Q1 | 0.004 | 0.002 | 1.73 | .08 | n.s. |
| IVH | decile | Q10 - Q1 | 0.005 | 0.002 | 2.42 | .02 | * |
| PVL | quartile | Q2 - Q1 | 0.001 | 0.001 | 1.11 | .27 | n.s. |
| PVL | quartile | Q3 - Q1 | 0.001 | 0.001 | 0.71 | .48 | n.s. |
| PVL | quartile | Q4 - Q1 | -0.001 | 0.002 | -0.90 | .37 | n.s. |
| PVL | quintile | Q2 - Q1 | 0.000 | 0.001 | -0.35 | .73 | n.s. |
| PVL | quintile | Q3 - Q1 | 0.000 | 0.001 | 0.29 | .77 | n.s. |
| PVL | quintile | Q4 - Q1 | 0.001 | 0.002 | 0.73 | .46 | n.s. |
| PVL | quintile | Q5 - Q1 | -0.003 | 0.002 | -1.61 | .11 | n.s. |
| PVL | decile | Q2 - Q1 | -0.001 | 0.001 | -1.20 | .23 | n.s. |
| PVL | decile | Q3 - Q1 | -0.002 | 0.001 | -1.70 | .09 | n.s. |
| PVL | decile | Q4 - Q1 | 0.000 | 0.002 | 0.08 | .93 | n.s. |
| PVL | decile | Q5 - Q1 | 0.001 | 0.002 | 0.32 | .75 | n.s. |
| PVL | decile | Q6 - Q1 | -0.001 | 0.002 | -0.56 | .58 | n.s. |
| PVL | decile | Q7 - Q1 | 0.002 | 0.002 | 0.76 | .45 | n.s. |

| | | | | | | | |
|---|---|---|---|---|---|---|---|
| PVL | decile | Q8 - Q1 | -0.001 | 0.002 | -0.36 | .71 | n.s. |
| PVL | decile | Q9 - Q1 | -0.007 | 0.003 | -2.28 | .02 | * |
| PVL | decile | Q10 - Q1 | -0.001 | 0.002 | -0.32 | .75 | n.s. |
| SEPS | quartile | Q2 - Q1 | 0.003 | 0.002 | 2.16 | .03 | * |
| SEPS | quartile | Q3 - Q1 | 0.002 | 0.002 | 1.07 | .28 | n.s. |
| SEPS | quartile | Q4 - Q1 | -0.001 | 0.002 | -0.61 | .54 | n.s. |
| SEPS | quintile | Q2 - Q1 | 0.003 | 0.002 | 1.72 | .09 | n.s. |
| SEPS | quintile | Q3 - Q1 | 0.003 | 0.002 | 1.58 | .11 | n.s. |
| SEPS | quintile | Q4 - Q1 | 0.001 | 0.002 | 0.55 | .58 | n.s. |
| SEPS | quintile | Q5 - Q1 | -0.002 | 0.002 | -1.22 | .22 | n.s. |
| SEPS | decile | Q2 - Q1 | 0.003 | 0.002 | 1.49 | .14 | n.s. |
| SEPS | decile | Q3 - Q1 | 0.004 | 0.002 | 1.75 | .08 | n.s. |
| SEPS | decile | Q4 - Q1 | 0.005 | 0.002 | 1.84 | .07 | n.s. |
| SEPS | decile | Q5 - Q1 | 0.004 | 0.002 | 1.44 | .15 | n.s. |
| SEPS | decile | Q6 - Q1 | 0.005 | 0.003 | 1.79 | .07 | n.s. |
| SEPS | decile | Q7 - Q1 | 0.000 | 0.002 | 0.15 | .88 | n.s. |

| | | | | | | | |
|---|---|---|---|---|---|---|---|
| SEPS | decile | Q8 - Q1 | 0.004 | 0.003 | 1.44 | .15 | n.s. |
| SEPS | decile | Q9 - Q1 | 0.003 | 0.003 | 1.35 | .18 | n.s. |
| SEPS | decile | Q10 - Q1 | -0.006 | 0.003 | -1.98 | .048 | * |
| NEC | quartile | Q2 - Q1 | 0.000 | 0.003 | 0.03 | .98 | n.s. |
| NEC | quartile | Q3 - Q1 | 0.002 | 0.003 | 0.58 | .56 | n.s. |
| NEC | quartile | Q4 - Q1 | 0.000 | 0.003 | 0.08 | .94 | n.s. |
| NEC | quintile | Q2 - Q1 | 0.000 | 0.003 | -0.10 | .92 | n.s. |
| NEC | quintile | Q3 - Q1 | 0.001 | 0.003 | 0.33 | .74 | n.s. |
| NEC | quintile | Q4 - Q1 | 0.000 | 0.003 | 0.08 | .93 | n.s. |
| NEC | quintile | Q5 - Q1 | 0.000 | 0.003 | 0.12 | .91 | n.s. |
| NEC | decile | Q2 - Q1 | -0.007 | 0.006 | -1.13 | .26 | n.s. |
| NEC | decile | Q3 - Q1 | -0.003 | 0.002 | -1.71 | .09 | n.s. |
| NEC | decile | Q4 - Q1 | -0.004 | 0.002 | -2.28 | .02 | * |
| NEC | decile | Q5 - Q1 | -0.004 | 0.002 | -1.77 | .08 | n.s. |
| NEC | decile | Q6 - Q1 | -0.001 | 0.002 | -0.69 | .49 | n.s. |
| NEC | decile | Q7 - Q1 | -0.003 | 0.002 | -1.32 | .19 | n.s. |

| | | | | | | | |
|---|---|---|---|---|---|---|---|
| NEC | decile | Q8 - Q1 | -0.003 | 0.003 | -1.24 | .21 | n.s. |
| NEC | decile | Q9 - Q1 | -0.004 | 0.002 | -2.04 | .04 | * |
| NEC | decile | Q10 - Q1 | -0.002 | 0.002 | -0.94 | .35 | n.s. |
| ROP | quartile | Q2 - Q1 | 0.000 | 0.001 | 0.19 | .85 | n.s. |
| ROP | quartile | Q3 - Q1 | 0.006 | 0.001 | 4.43 | <.001 | *** |
| ROP | quartile | Q4 - Q1 | 0.009 | 0.001 | 6.61 | <.001 | *** |
| ROP | quintile | Q2 - Q1 | 0.001 | 0.002 | 0.52 | .60 | n.s. |
| ROP | quintile | Q3 - Q1 | 0.000 | 0.002 | 0.02 | .99 | n.s. |
| ROP | quintile | Q4 - Q1 | 0.008 | 0.001 | 5.87 | <.001 | *** |
| ROP | quintile | Q5 - Q1 | 0.011 | 0.002 | 6.56 | <.001 | *** |
| ROP | decile | Q2 - Q1 | -0.001 | 0.002 | -0.48 | .63 | n.s. |
| ROP | decile | Q3 - Q1 | -0.001 | 0.002 | -0.38 | .71 | n.s. |
| ROP | decile | Q4 - Q1 | 0.001 | 0.002 | 0.55 | .58 | n.s. |
| ROP | decile | Q5 - Q1 | -0.001 | 0.003 | -0.26 | .80 | n.s. |
| ROP | decile | Q6 - Q1 | 0.000 | 0.002 | -0.14 | .89 | n.s. |
| ROP | decile | Q7 - Q1 | 0.009 | 0.002 | 4.69 | <.001 | *** |

| | | | | | | | |
|---|---|---|---|---|---|---|---|
| ROP | decile | Q8 - Q1 | 0.006 | 0.002 | 3.07 | .002 | ** |
| ROP | decile | Q9 - Q1 | 0.008 | 0.002 | 3.33 | .001 | ** |
| ROP | decile | Q10 - Q1 | 0.012 | 0.002 | 5.53 | <.001 | *** |
| Death | quartile | Q2 - Q1 | -0.001 | 0.001 | -0.58 | .56 | n.s. |
| Death | quartile | Q3 - Q1 | 0.003 | 0.001 | 2.09 | .04 | * |
| Death | quartile | Q4 - Q1 | 0.010 | 0.001 | 8.31 | <.001 | *** |
| Death | quintile | Q2 - Q1 | 0.000 | 0.002 | 0.01 | .99 | n.s. |
| Death | quintile | Q3 - Q1 | 0.001 | 0.001 | 0.84 | .40 | n.s. |
| Death | quintile | Q4 - Q1 | 0.006 | 0.002 | 3.71 | <.001 | *** |
| Death | quintile | Q5 - Q1 | 0.011 | 0.001 | 8.66 | <.001 | *** |
| Death | decile | Q2 - Q1 | 0.002 | 0.002 | 0.82 | .41 | n.s. |
| Death | decile | Q3 - Q1 | 0.000 | 0.002 | 0.14 | .89 | n.s. |
| Death | decile | Q4 - Q1 | 0.001 | 0.002 | 0.61 | .54 | n.s. |
| Death | decile | Q5 - Q1 | 0.002 | 0.002 | 0.90 | .37 | n.s. |
| Death | decile | Q6 - Q1 | 0.002 | 0.002 | 1.11 | .27 | n.s. |
| Death | decile | Q7 - Q1 | 0.003 | 0.002 | 1.35 | .18 | n.s. |

| | | | | | | | |
|---|---|---|---|---|---|---|---|
| Death | decile | Q8 - Q1 | 0.010 | 0.002 | 4.71 | <.001 | *** |
| Death | decile | Q9 - Q1 | 0.009 | 0.002 | 4.54 | <.001 | *** |
| Death | decile | Q10 - Q1 | 0.016 | 0.002 | 8.33 | <.001 | *** |
| NHI | quartile | Q2 - Q1 | 0.017 | 0.015 | 1.11 | .27 | n.s. |
| NHI | quartile | Q3 - Q1 | 0.036 | 0.012 | 2.97 | .003 | ** |
| NHI | quartile | Q4 - Q1 | 0.092 | 0.011 | 8.08 | <.001 | *** |
| NHI | quintile | Q2 - Q1 | 0.018 | 0.017 | 1.03 | .30 | n.s. |
| NHI | quintile | Q3 - Q1 | 0.016 | 0.016 | 1.00 | .32 | n.s. |
| NHI | quintile | Q4 - Q1 | 0.059 | 0.014 | 4.27 | <.001 | *** |
| NHI | quintile | Q5 - Q1 | 0.093 | 0.013 | 7.19 | <.001 | *** |
| NHI | decile | Q2 - Q1 | 0.018 | 0.023 | 0.79 | .43 | n.s. |
| NHI | decile | Q3 - Q1 | 0.013 | 0.028 | 0.47 | .64 | n.s. |
| NHI | decile | Q4 - Q1 | 0.040 | 0.026 | 1.55 | .12 | n.s. |
| NHI | decile | Q5 - Q1 | 0.019 | 0.027 | 0.72 | .47 | n.s. |
| NHI | decile | Q6 - Q1 | 0.030 | 0.023 | 1.30 | .19 | n.s. |
| NHI | decile | Q7 - Q1 | 0.053 | 0.024 | 2.23 | .03 | * |

| | | | | | | | |
|---|---|---|---|---|---|---|---|
| NHI | decile | Q8 - Q1 | 0.083 | 0.022 | 3.73 | <.001 | *** |
| NHI | decile | Q9 - Q1 | 0.088 | 0.022 | 3.99 | <.001 | *** |
| NHI | decile | Q10 - Q1 | 0.116 | 0.022 | 5.30 | <.001 | *** |

**Table S8**. Variable importance

| Rank | Variable | Variable importance |
|---|---|---|
| 1 | GA at PPROM (weeks) | 0.350 |
| 2 | Oligohydramnios | 0.189 |
| 3 | Maternal age | 0.133 |
| 4 | Size for GA (SGA/AGA/LGA) | 0.088 |
| 5 | Multiple gestation | 0.058 |
| 6 | In Vitro Fertilization pregnancy | 0.053 |
| 7 | Fetal sex | 0.045 |
| 8 | Primiparity | 0.039 |
| 9 | Gestational/Overt diabetes mellitus | 0.038 |
| 10 | Chronic/Pregnancy-induced hypertension | 0.009 |

**Table S9**. Best linear projection (BLP) results. BLP was performed by regressing doubly robust (DR) scores on all covariates. The estimated coefficients represent the association between each covariate and variation in individual treatment effects (ITEs), conditional on the other covariates. n.s., not significant; *** p < .001; **p < .01; *p < .05

| Variable | blp_coef | blp_se | blp_t | blp_p | ci_lower | ci_upper | significance |
|---|---|---|---|---|---|---|---|
| (Intercept) | 0.251 | 0.071 | 3.543 | <.001 | 0.112 | 0.390 | |
| Fetal sex | -0.013 | 0.009 | -1.419 | .16 | -0.030 | 0.005 | |
| Multiple gestation | -0.013 | 0.011 | -1.221 | .22 | -0.034 | 0.008 | |
| In vitro fertilization | -0.020 | 0.012 | -1.660 | .10 | -0.043 | 0.004 | |
| Gestational/Overt Diabetes Mellitus | -0.015 | 0.014 | -1.126 | .26 | -0.042 | 0.011 | |
| Chronic/ Pregnancy-induced hypertension | 0.057 | 0.019 | 2.993 | .003 | 0.020 | 0.095 | ** |
| Oligohydramnios | -0.042 | 0.011 | -3.745 | <.001 | -0.064 | -0.020 | *** |
| Primiparity | -0.003 | 0.009 | -0.323 | .75 | -0.021 | 0.015 | |
| Size for gestational age | 0.059 | 0.031 | 1.914 | .06 | -0.001 | 0.120 | |
| Maternal age | 0.000 | 0.001 | -0.357 | .72 | -0.002 | 0.002 | |
| Gestational age at PPROM (weeks) | -0.009 | 0.001 | -6.972 | <.001 | -0.012 | -0.007 | *** |

**Table S10.** Predicted Neonatal Health Index (NHI) at selected latency periods derived from restricted cubic spline analysis with cluster-robust variance estimation (sensitivity analysis). Values are presented as predicted NHI (95% confidence interval); NHI ranges from 0 (worst) to 8 (best). A dash(—) indicates that the selected timepoint exceeded the 99th percentile of observed latency within that GA group.

| Latency period (days) | Full cohort model (GA-adjusted, n=6,872) | Subgroup-specific models | | | |
|---|---|---|---|---|---|
| | | <24 weeks (n=1,270) | 24–25+6 weeks (n=1,238) | 26–27+6 weeks (n=1,423) | ≥28 weeks (n=2,941) |
| | Predicted NHI (95% CI) | Predicted NHI (95% CI) | Predicted NHI (95% CI) | Predicted NHI (95% CI) | Predicted NHI (95% CI) |
| 0 | 5.19 [5.09-5.28] | 1.36 [1.10-1.66] | 3.81 [3.52-4.08] | 5.86 [5.67-6.06] | 7.28 [7.21-7.36] |
| 1 | 5.44 [5.36-5.53] | 1.60 [1.36-1.85] | 4.08 [3.88-4.27] | 6.24 [6.10-6.38] | 7.37 [7.30-7.42] |
| 2 | 5.66 [5.58-5.75] | 1.87 [1.63-2.10] | 4.33 [4.10-4.56] | 6.52 [6.34-6.68] | 7.45 [7.37-7.50] |
| 3 | 5.77 [5.69-5.86] | 2.14 [1.89-2.38] | 4.58 [4.30-4.85] | 6.68 [6.50-6.85] | 7.50 [7.42-7.56] |
| 4 | 5.90 [5.81-5.98] | 2.40 [2.12-2.67] | 4.81 [4.50-5.12] | 6.78 [6.61-6.92] | 7.54 [7.47-7.61] |
| 5 | 6.00 [5.91-6.08] | 2.63 [2.33-2.94] | 5.03 [4.71-5.34] | 6.81 [6.66-6.94] | 7.57 [7.50-7.64] |
| 6 | 6.07 [5.99-6.15] | 2.84 [2.51-3.17] | 5.23 [4.92-5.52] | 6.83 [6.68-6.96] | 7.60 [7.53-7.66] |
| 7 | 6.12 [6.04-6.19] | 3.02 [2.69-3.37] | 5.41 [5.14-5.68] | 6.86 [6.69-7.00] | 7.61 [7.55-7.68] |
| 14 | 6.51 [6.44-6.59] | 3.69 [3.44-4.01] | 6.30 [6.02-6.57] | 7.17 [7.01-7.30] | 7.70 [7.56-7.83] |
| 21 | 6.82 [6.74-6.89] | 3.98 [3.65-4.35] | 6.74 [6.54-6.95] | 7.50 [7.36-7.64] | — |
| 28 | 7.06 [7.00-7.13] | 4.30 [3.99-4.63] | 7.04 [6.77-7.26] | 7.71 [7.55-7.84] | — |
| 35 | 7.28 [7.23-7.33] | 4.61 [4.32-4.91] | 7.26 [6.88-7.54] | — | — |

| | | | | | |
|---|---|---|---|---|---|
| 42 | 7.44 [7.40-7.49] | 4.91 [4.56-5.25] | — | — | — |
| 49 | 7.57 [7.53-7.61] | 5.15 [4.78-5.51] | — | — | — |
| 56 | 7.68 [7.64-7.71] | 5.38 [5.02-5.72] | — | — | — |
| 63 | 7.76 [7.72-7.79] | 5.57 [5.23-5.92] | — | — | — |
| 70 | 7.82 [7.79-7.85] | 5.74 [5.34-6.09] | — | — | — |
| 77 | — | 5.90 [5.44-6.28] | — | — | — |
| 84 | — | 6.07 [5.49-6.52] | — | — | — |
| 91 | — | 6.21 [5.56-6.74] | — | — | — |
| 97 | — | 6.33 [5.59-6.90] | — | — | — |

**Table S11.** Estimated average partial effects (APE) and model calibration results of the Neonatal Health Index (NHI) in the sensitivity analysis

| grf | | | | | | | EconML | |
|---|---|---|---|---|---|---|---|---|
| Outcome | APE | p | β_ATE | p | β_ITE | p | APE | p |
| PH | 0.0031 | <.001 | 1.32 | <.001 | -30.78 | 1.00 | 0.0036 | .13 |
| BPD | 0.0119 | <.001 | 1.01 | <.001 | -0.78 | .66 | 0.0131 | <.001 |
| IVH | 0.0060 | <.001 | 0.88 | <.001 | 2.19 | <.001 | 0.0070 | <.001 |

| | | | | | | | | |
|---|---|---|---|---|---|---|---|---|
| PVL | 0.0047 | <.001 | 0.49 | <.001 | -206.40 | 1.00 | 0.0042 | .02 |
| SEPS | 0.0096 | <.001 | 1.05 | <.001 | -36.41 | 1.00 | 0.0097 | <.001 |
| NEC | 0.0015 | .09 | 1.47 | <.001 | -18.48 | 1.00 | 0.0029 | .10 |
| ROP | 0.0075 | <.001 | 0.85 | <.001 | 1.32 | <.001 | 0.0088 | <.001 |
| Death | 0.0073 | <.001 | 0.90 | <.001 | 1.13 | <.001 | 0.0077 | .13 |
| NHI | 0.0866 | <.001 | 0.90 | <.001 | 1.29 | <.001 | 0.0932 | <.001 |

**Table S12.** Group average treatment effects (GATEs) for quartile-, quintile-, and decile-based stratifications of predicted individual treatment effects (ITEs) in sensitivity analysis. P-values test the null hypothesis that the GATE equals zero using two-sided Wald tests. n.s., not significant; *** p < .001; ** p < .01; * p < .05.

| Quantile type | Quantile group | CI_low | CI_up | GATE_est | GATE_se | GATE_p | Significance |
|---|---|---|---|---|---|---|---|
| quartile | Q1 | 0.0595 | 0.0738 | 0.0319 | 0.0121 | .008 | ** |
| quartile | Q2 | 0.0738 | 0.0854 | 0.0639 | 0.0111 | <.001 | *** |
| quartile | Q3 | 0.0854 | 0.1331 | 0.0907 | 0.0081 | <.001 | *** |
| quartile | Q4 | 0.1331 | 0.1688 | 0.1601 | 0.0084 | <.001 | *** |
| quintile | Q1 | 0.0595 | 0.0715 | 0.0315 | 0.0129 | .01 | * |
| quintile | Q2 | 0.0715 | 0.0795 | 0.0604 | 0.0140 | <.001 | *** |
| quintile | Q3 | 0.0795 | 0.0933 | 0.0695 | 0.0098 | <.001 | *** |

| | | | | | | | |
|---|---|---|---|---|---|---|---|
| quintile | Q4 | 0.0933 | 0.1442 | 0.1110 | 0.0095 | <.001 | *** |
| quintile | Q5 | 0.1442 | 0.1688 | 0.1607 | 0.0095 | <.001 | *** |
| decile | Q1 | 0.0595 | 0.0673 | 0.0214 | 0.0190 | .26 | n.s. |
| decile | Q2 | 0.0673 | 0.0715 | 0.0417 | 0.0176 | .02 | * |
| decile | Q3 | 0.0715 | 0.0753 | 0.0631 | 0.0196 | .001 | ** |
| decile | Q4 | 0.0753 | 0.0795 | 0.0577 | 0.0198 | .004 | ** |
| decile | Q5 | 0.0795 | 0.0854 | 0.0557 | 0.0156 | <.001 | *** |
| decile | Q6 | 0.0854 | 0.0933 | 0.0833 | 0.0118 | <.001 | *** |
| decile | Q7 | 0.0933 | 0.1260 | 0.0905 | 0.0144 | <.001 | *** |
| decile | Q8 | 0.1260 | 0.1442 | 0.1316 | 0.0124 | <.001 | *** |
| decile | Q9 | 0.1442 | 0.1546 | 0.1459 | 0.0135 | <.001 | *** |
| decile | Q10 | 0.1546 | 0.1688 | 0.1755 | 0.0133 | <.001 | *** |

**Table S13.** Pairwise contrasts in group average treatment effects (GATEs) comparing Q1 with higher quantile groups in sensitivity analysis. Contrasts were calculated as the difference in estimated GATE between each subgroup and Q1 using Wald tests. n.s., not significant; *** $p < .001$; ** $p < .01$; * $p < .05$.

| Quantile type | contrast | diff | se | z | p | Significance |
|---|---|---|---|---|---|---|

| | | | | | | |
|---|---|---|---|---|---|---|
| quartile | Q2 - Q1 | 0.032 | 0.016 | 1.94 | .052 | n.s. |
| quartile | Q3 - Q1 | 0.059 | 0.015 | 4.02 | <.001 | *** |
| quartile | Q4 - Q1 | 0.128 | 0.015 | 8.69 | <.001 | *** |
| quintile | Q2 - Q1 | 0.029 | 0.019 | 1.52 | .13 | n.s. |
| quintile | Q3 - Q1 | 0.038 | 0.016 | 2.34 | .02 | * |
| quintile | Q4 - Q1 | 0.079 | 0.016 | 4.95 | <.001 | *** |
| quintile | Q5 - Q1 | 0.129 | 0.016 | 8.05 | <.001 | *** |
| decile | Q2 - Q1 | 0.020 | 0.026 | 0.79 | .43 | n.s. |
| decile | Q3 - Q1 | 0.042 | 0.027 | 1.52 | .13 | n.s. |
| decile | Q4 - Q1 | 0.036 | 0.027 | 1.32 | .19 | n.s. |
| decile | Q5 - Q1 | 0.034 | 0.025 | 1.40 | .16 | n.s. |
| decile | Q6 - Q1 | 0.062 | 0.022 | 2.77 | .006 | ** |
| decile | Q7 - Q1 | 0.069 | 0.024 | 2.90 | .004 | ** |
| decile | Q8 - Q1 | 0.110 | 0.023 | 4.86 | <.001 | *** |
| decile | Q9 - Q1 | 0.125 | 0.023 | 5.34 | <.001 | *** |
| decile | Q10 - Q1 | 0.154 | 0.023 | 6.64 | <.001 | *** |

**Table S14.** Covariate comparison between the low (Q1) and high-benefit (Q5) group in GATE test in the sensitivity analysis

| Variable | | Q1 (N=1280) | Q5 (N=1280) | p | Standardized mean difference |
|---|---|---|---|---|---|
| Maternal age, mean(SD) | | 32.90 (3.54) | 33.27 (4.13) | .01 | 0.096 |
| Gestational age at PPROM (weeks), mean(SD) | | 29.84 (1.48) | 22.85 (1.20) | <.001 | -5.195 |
| Multiple gestation, n(%) | | 1060 (82.8%) | 427 (33.4%) | <.001 | -0.495 |
| In vitro fertilization, n(%) | | 717 (56.0%) | 396 (30.9%) | <.001 | -0.251 |
| Gestational or overt Diabetes Mellitus, n(%) | | 182 (14.2%) | 73 (5.7%) | <.001 | -0.085 |
| Chronic or pregnancy-induced hypertension, n(%) | | 86 (6.7%) | 48 (3.8%) | .001 | -0.030 |
| Oligohydramnios, n(%) | | 52 (4.1%) | 270 (21.1%) | <.001 | 0.170 |
| Primiparity, n(%) | | 1174 (91.7%) | 780 (60.9%) | <.001 | -0.308 |
| Fetal sex (Male), n(%) | | 428 (33.4%) | 632 (49.4%) | <.001 | -0.159 |
| Size for gestational age, n(%) | SGA | 160 (12.5%) | 40 (3.1%) | <.001 | -0.094 |
| | AGA | 1116 (87.2%) | 1202 (93.9%) | | 0.067 |
| | LGA | 4 (0.3%) | 38 (3.0%) | | 0.027 |

**Table S15**. Variable importance obtained in the sensitivity analysis

| Rank | Variable | Variable importance |
|---|---|---|
| 1 | Gestational age at PPROM (weeks) | 0.623 |
| 2 | Maternal age | 0.129 |
| 3 | Oligohydramnios | 0.081 |
| 4 | Fetal sex | 0.056 |
| 5 | Primiparity | 0.042 |
| 6 | In vitro fertilization | 0.036 |
| 7 | Multiple gestation | 0.029 |
| 8 | Gestational or overt Diabetes Mellitus | 0.005 |
| 9 | Chronic or pregnancy-induced hypertension | 0 |
| 10 | Size for gestational age | 0 |

**Table S16.** Best linear projection (BLP) results in the sensitivity analysis. BLP was performed by regressing doubly robust (DR) scores on all covariates. The estimated coefficients represent the association between each covariate and variation in individual treatment effects (ITEs), conditional on the other covariates. n.s., not significant; *** p < .001; **p < .01; *p < .05

| Variable | blp_coef | blp_se | blp_t | blp_p | ci_lower | ci_upper | significance |
|---|---|---|---|---|---|---|---|
| (Intercept) | 0.572 | 0.082 | 6.952 | <.001 | 0.410 | 0.733 | *** |

| | | | | | | | |
|---|---|---|---|---|---|---|---|
| Fetal sex | -0.012 | 0.010 | -1.161 | .25 | -0.031 | 0.008 | |
| Multiple gestation | -0.003 | 0.011 | -0.271 | .79 | -0.026 | 0.019 | |
| In vitro fertilization | -0.015 | 0.013 | -1.176 | .24 | -0.041 | 0.010 | |
| Gestational or overt Diabetes Mellitus | -0.001 | 0.014 | -0.091 | .93 | -0.028 | 0.026 | |
| Chronic or pregnancy-induced hypertension | 0.053 | 0.020 | 2.584 | .01 | 0.013 | 0.093 | ** |
| Oligohydramnio s | -0.042 | 0.014 | -3.061 | .002 | -0.069 | -0.015 | ** |
| Primiparity | -0.014 | 0.010 | -1.367 | .17 | -0.034 | 0.006 | |
| Size for gestational age | 0.008 | 0.031 | 0.268 | .79 | -0.053 | 0.070 | |
| Maternal age | -0.001 | 0.001 | -0.778 | .44 | -0.003 | 0.001 | |
| Gestational age at PPROM (weeks) | -0.017 | 0.002 | -8.581 | <.001 | -0.020 | -0.013 | *** |

**Figure S1. Flowchart of study population selection**

A total of 21,400 infants registered in the Korean Neonatal Network (KNN) were screened for eligibility. Infants were excluded if they were not coded as PPROM (neonates with no PROM(n = 13,842), had unknown PROM status (n = 183), or had missing information on PROM onset or delivery time (n = 312) (total excluded: n = 14,337). Of the remaining 7,063 eligible PPROM cases, 2 were further excluded due to missing PROM onset date or time. 4 neonates were excluded for implausible latency periods exceeding 120 days. The final analytic cohort comprised 7,057 neonates with confirmed PPROM.

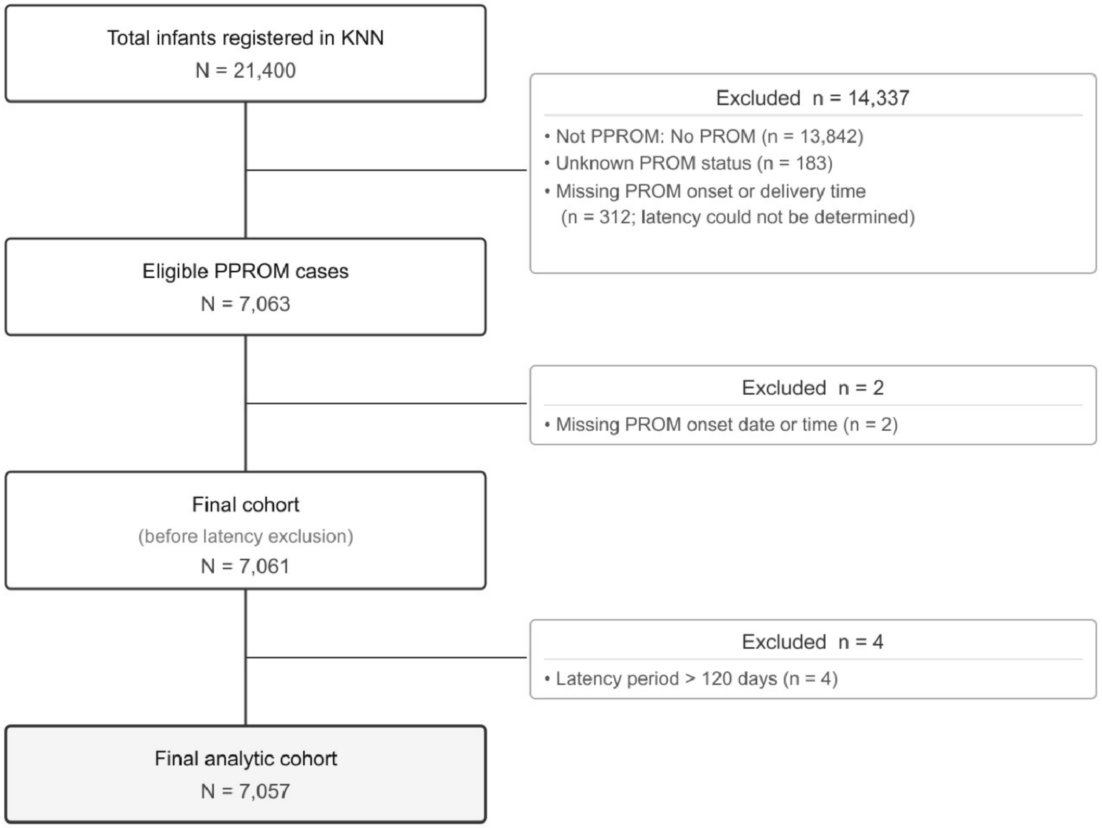

**Figure S2. Group average treatment effects (GATEs) for quartile- and decile-based stratifications of predicted individual treatment effects (ITEs) of NHI.** The estimated GATE with 95% confidence intervals is shown for each quantile group. Asterisks indicate statistical significance of the GATE estimates, and horizontal bars denote pairwise comparisons between Q1 and the other quintiles. Statistical significance of the GATE estimates and pairwise comparisons between Q1 and the other quantile groups were conducted using Wald tests. n.s., not significant; *** p < .001; ** p < .01; * p < .05.

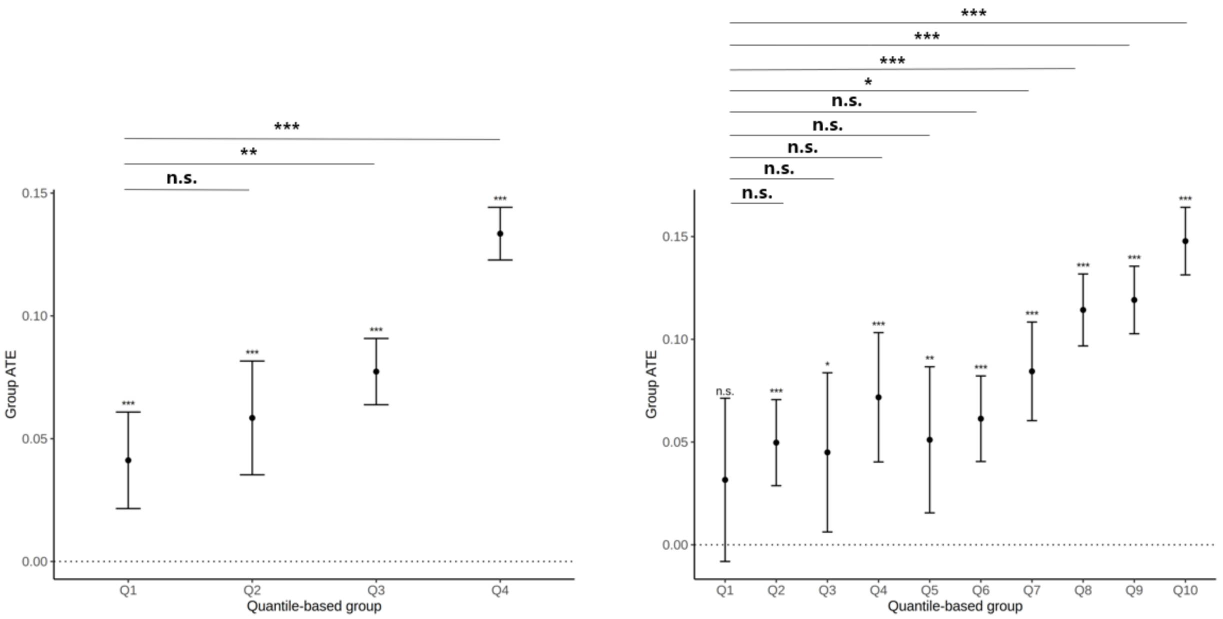

**Figure S3. Group average treatment effects (GATEs) for quartile- and decile-based stratifications of predicted individual treatment effects (ITEs) of morbidities and Death used to construct the NHI.** The estimated GATE with 95% confidence intervals is shown for each quantile group. Asterisks indicate statistical significance of the GATE estimates assessed using Wald tests. n.s., not significant; *** p < .001; ** p < .01; * p < .05.

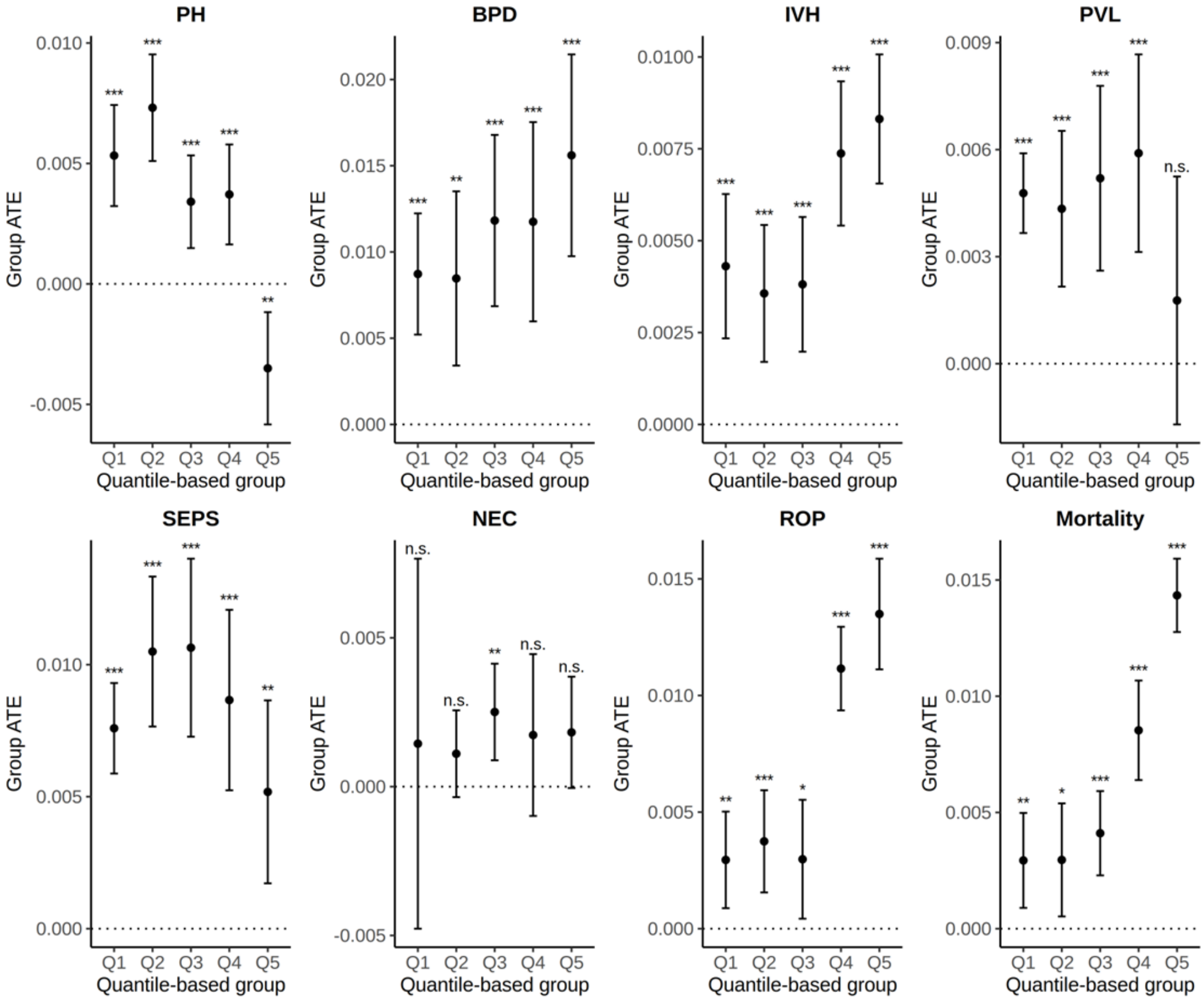

**Figure S4.** Variable importance of the secondary outcomes comprising the NHI

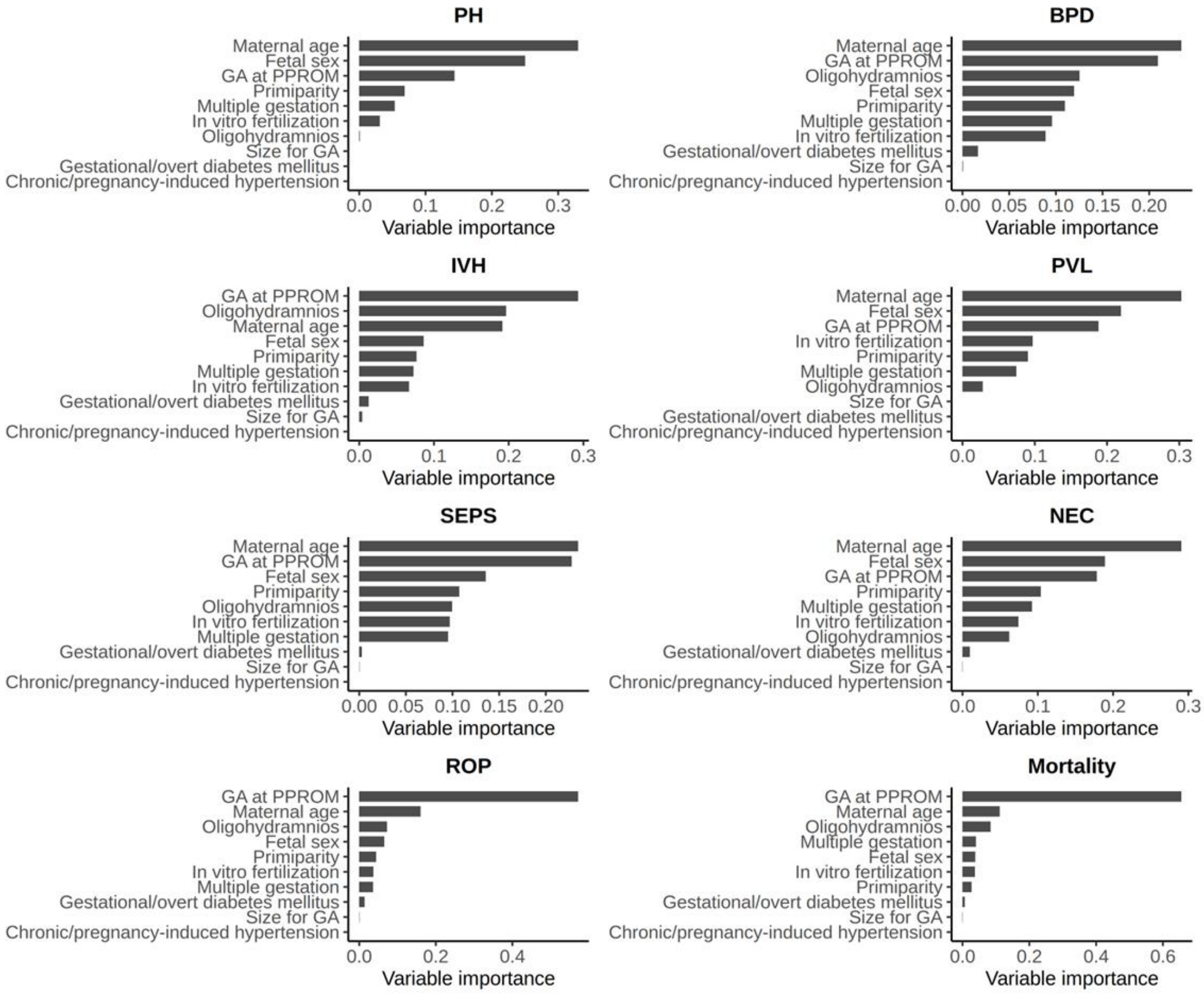

**Figure S5. CATE interpreter trees derived from individual treatment effects (ITEs) estimated from the causal forest model for the NHI in EconML.**
Trees were constructed from the ITE estimates to summarize heterogeneous treatment effects across subgroups. Different hyperparameter combinations were explored in the CATE interpreter tree, specifically varying the maximum tree depth (max_depth ∈ {2, 3, 4}) and the minimum leaf node size (min_samples_leaf ∈ {500, 1000, 1500}). Trees with greater depths were not considered to preserve interpretability. Larger minimum leaf node sizes were also excluded as they resulted in trivial splits dominated solely by GA at PPROM. **A**, Maximum depth = 2, minimum leaf node size = 500. **B**, Maximum depth = 3, minimum leaf node size = 1000. **C,** Maximum depth = 4, minimum leaf node size = 1500.

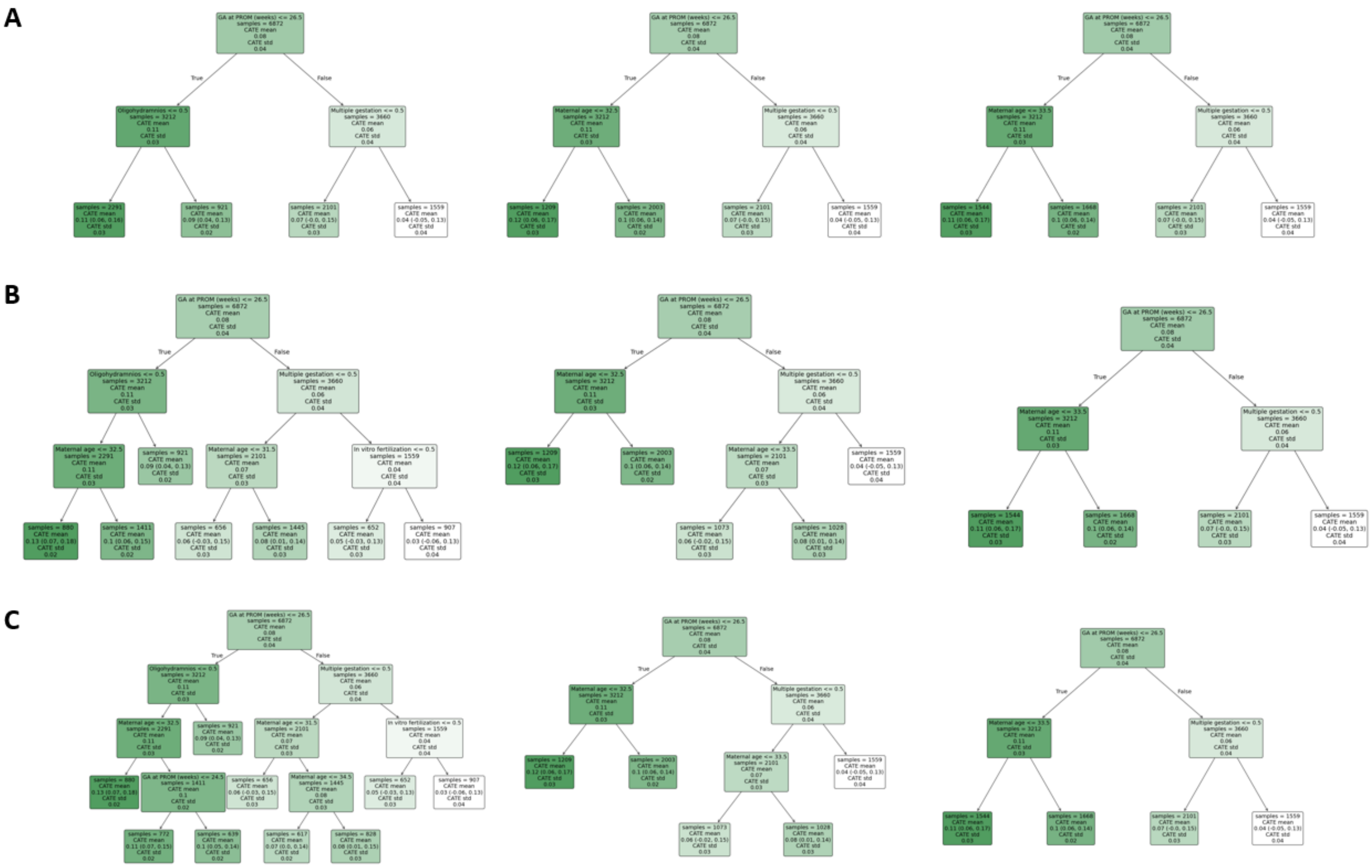

**Figure S6. Identification of variables contributing to treatment effect heterogeneity for the NHI in a sensitivity analysis restricting the sample to patients with latency period ≤ 30 days (N = 6,575).**

**A**, Quintile-based group average treatment effect (GATE) test. Patients were stratified into quintiles according to predicted individual treatment effects (ITEs), and the estimated GATE with 95% confidence intervals is shown for each group. Asterisks indicate statistical significance of the GATE estimates, and horizontal bars denote pairwise comparisons between Q1 and the other quintiles. **B**, Variable importance from the causal forest fitted in *grf*. *** $p < .001$; **$p < .01$; *$p < .05$

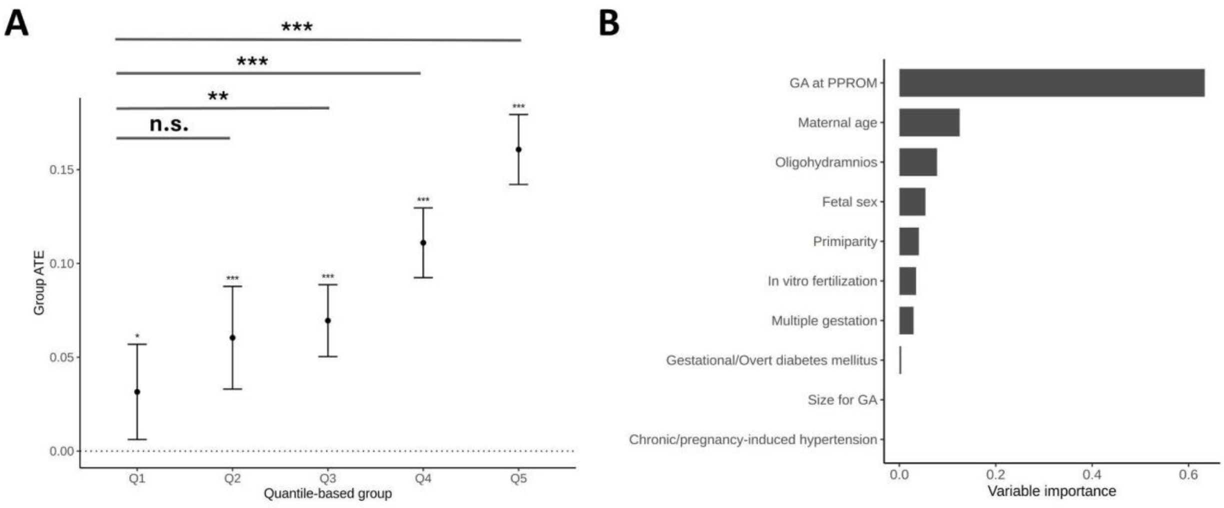
